# A systematic review of observational methods used to quantify personal protective behaviours among members of the public during the COVID-19 pandemic, and the concordance between observational and self-report measures in infectious disease health protection

**DOI:** 10.1101/2021.12.22.21268226

**Authors:** Rachel Davies, Fiona Mowbray, Alex F Martin, Louise E Smith, G James Rubin

## Abstract

**Objectives:** To assess the quantity and quality of studies using an observational measure of behaviour during the COVID-19 pandemic, and to narratively describe the association between self-report and observational data for behaviours relevant to controlling an infectious disease outbreak.

**Design:** Systematic review and narrative synthesis of observational studies.

**Data sources:** We searched Medline, Embase, PsychInfo, Publons, Scopus and the Public Health England behavioural science LitRep database from inception to 17^th^ September 2021 for relevant studies.

**Study selection:** We included studies which collected observational data of at least one of three health protective behaviours (hand hygiene, face covering use and maintaining physical distance from others (‘social distancing’)) during the COVID-19 pandemic. Studies where observational data were compared to self-report data in relation to any infectious disease were also included.

**Data extraction and synthesis:** We evaluated the quality of studies using the NIH quality assessment scale for observational studies, extracted data on sample size, setting and adherence to health protective behaviours, and synthesized results narratively.

**Results:** Of 27,279 published papers on COVID-19 relevant health protective behaviours that included one or more terms relating to hand hygiene, face covering and social distancing, we identified 48 studies that included an objective observational measure. Of these, 35 assessed face covering use, 17 assessed hand hygiene behaviour and seven assessed physical distancing. The general quality of these studies was good. When expanding the search to all infectious diseases, we included 21 studies that compared observational versus self-report data. These almost exclusively studied hand hygiene. The difference in outcomes was striking, with self-report over-estimating observed adherence by up to a factor of five in some settings. In only four papers did self-report match observational data in any domains.

**Conclusions:** Despite their importance in controlling the pandemic, we found remarkably few studies assessing protective behaviours by observation, rather than self-report, though these studies tended to be of reasonably good quality. Observed adherence tends to be substantially lower than estimates obtained via self-report. Accurate assessment of levels of personal protective behaviour, and evaluation of interventions to increase this, would benefit from the use of observational methods.

## Background

Throughout the COVID-19 pandemic, members of the public have been urged to engage in a set of behaviours intended to reduce transmission of the SARS-CoV-2 virus. These have included recommendations to practice frequent hand hygiene, avoid close contact with other people (‘social distancing’) and to wear a face covering to prevent spread through respiratory droplets.^1, 2^ Although these interventions have been shown to be effective in reducing the transmission of SARS-CoV-2,^3^ none of the interventions will work if people do not adhere to them or understand the messaging on when they should adhere.^4^

To date, public engagement with recommended behaviours has primarily been monitored by behavioural scientists, public health agencies and national governments though the use of self-report questionnaires. Using self-report methods to collect data has many benefits. Self-reported data can be quick, easy and relatively inexpensive to obtain from large numbers of participants. The association between self-reported behaviour and other self-reported variables is also relatively straightforward to examine. For many outcomes, self-report can be a good proxy measure for actual behaviour^5^. For example, self-report can be a useful way to assess whether someone has been vaccinated or not^6^. For other behaviours, self-report may be less valid^7^. This may be particularly true for frequently performed behaviours that are difficult to remember (e.g. frequency of handwashing in the past 24 hours) or that would be socially undesirable to admit (e.g. breaking legally enforceable rules around self-isolation)^8, 9, 10^.

In the context of COVID-19, regularly collected measures of behaviour that do not rely on self-report are rare. Notable exceptions include: mobility data based on mobile phone locations^11^; footfall data in city centres^12^; and official statistics on vaccine uptake based on electronic records^13^. Most of these examples relate to where people are located or whether they engage with health services.

There are fewer regularly collected data based on direct observation quantifying whether and how people engage with COVID-19 protective behaviours. To assist public health agencies in considering whether to collect more observational data, we conducted a systematic review of the use of observational measures of COVID-19 relevant behaviours. We focussed on studies that directly observed the performance of protective behaviours, excluding measures of mobility or location. Our aims were to assess: 1) the quantity of observational studies conducted during COVID-19; 2) the quality of these studies; and 3) the association between self-report and observational data. While we only included COVID-19 related studies for aims one and two, given a lack of data found during screening, we expanded our inclusion criteria for aim three and included studies relating to any infectious disease outbreak.

## Methods

### Protocol and registration

This review follows the PRISMA framework and is registered with PROSPERO registration number CRD42021261360. The study protocol is available from: https://www.crd.york.ac.uk/prospero/display_record.php?ID=CRD42021261360

### Search strategies

For aims one and two, we searched the following electronic databases from inception to 17^th^ September 2021: Medline, Embase, PsycInfo, Publons, Scopus, and the Public Health England (PHE) behavioural science COVID-19 Literature Repository database (BSIU LitRep Database. Google Docs. Available from:https://docs.google.com/spreadsheets/d/1qfR4NgnD5hTAS8KriPaXYhLu1s7fpZJDq8EIXQY0ZEs/edit#gid=369408275 (Accessed November 2021)). Databases were searched for articles containing MeSH terms or keywords relating to COVID-19 (e.g. “SARS-CoV-2”, “novel coronavirus”), hand hygiene, physical distancing, or face coverings (e.g. “hand washing”, “face mask”, “physical distancing”) and an observational method (e.g. “observational study”, “videorecording”). Full details of our search strategies are available in Supplementary material.

For aim three, we searched Medline, Embase and PsycInfo from inception to 17^th^ September 2021. These were searched for articles containing MeSH terms or keywords relating to hand hygiene, physical distancing, face covering and direct observation. We did not include specific search terms for infectious disease as this was already the focus of the majority of papers investigating the three relevant behaviours.

For both search strategies we examined the reference sections of any pertinent studies and reviews for further references.

### Eligibility criteria

For aims one and two, we included studies that were published in English since January 2020, contained an observational measure of hand hygiene, physical distancing or face covering use in relation to COVID-19, assessed these behaviours among the general public or healthcare workers, and contained original data. We excluded studies that contained only location-based data, for example, mobile phone data that measured where in space people were located (rather than what they were doing). We also excluded the use of used crowd density measurements where physical distancing of individuals within the crowd could not be determined. Studies were also excluded if they recorded impressionistic perceptions of behaviour rather than using a systematic method such as using unsystematic sampling methods or retrospective methods based on recall of behaviours.

For aim three, we included studies published in English (no date restrictions), that related to infectious disease control for any pathogen and that contained an observational measure of one or more of our defined behaviours compared to a self-report measure. We excluded studies that contained only self-report or observational data.

### Study selection

Titles and abstracts were independently double screened by two separate reviewers (RD screened all citations, FM screened half the citations and AFM screened the other half) using Sysrev Software to identify potentially eligible studies and record decisions. Full texts were then independently double screened (RD screened all citations, FM screened half the citations and AFM screened the other half), with any uncertainties resolved through discussion.

### Data extraction, items and risk of bias

Two reviewers (RD, FM) extracted data from included studies. Study and participant characteristics were noted, including study design, sample size, number of opportunities for specified behaviours, location of observation, population characteristics and prevalence of adherence. Where needed, further details were sought by contacting study authors. For aims one and two, where papers contained a pre and post COVID-19 data collection period, only data collected during the COVID-19 pandemic were included in the narrative synthesis.

Studies were assessed for quality using the National Institutes of Health (NIH) Quality Assessment Tool for Observational Cohort and Cross Sectional Studies^14^.

## Results

27,279 published papers were identified that included terms relating to COVID-19, and one or more terms relating to hand hygiene, face covering and social distancing. When the term ‘observational’ and related terms were added, 2589 papers were identified and these were screened for aims one and two, from which 105 were selected as potentially relevant to the review. Of these, 57 were excluded. A total of 48 studies met the inclusion criteria (See supplementary material). For aim three we screened 3,331 papers, from which 133 were deemed potentially relevant following abstract screening. Of these, 21 were included in the review.

### Aim one: Quantity of studies using observational measures

We included 48^15–62^ studies containing an observation component during the COVID-19 pandemic. In total these included at least 116,169 participants and at least 36,060,422 behavioural observational events.

Of the included studies, 39 used direct observers, one used an automated measurement to assess hand hygiene, five used video observations and three used mixed methods including; observation supplemented with a survey, observation supplemented with media data and in-person observation plus automated technology.

Of the 48 included studies, 35 looked at wearing a face covering (five in healthcare workers, 30 in the general public), 17 studies looking at hand hygiene (12 in healthcare workers, five in the general public), and seven looked at physical distancing (one in healthcare workers and six in the general public).

Six studies contained an interventional component intended to improve adherence.

Studies had been conducted in Asia (n=18), North America (n=15), Europe (14), Africa (n=2), and Australia (n=1).

The most common setting for observation was in hospitals (n=20), in stores or shopping centres (n=12), on public streets (n=11), on public transport (n=7), in parks (n=4), high schools or universities (n=3), community healthcare (n=2) and residential care homes (n=1).

For papers for which it could be determined (N=43), sample size varied between 41 and 17,200 (median = 780).

The median number of behavioural observation events for each study was 1,020, with a minimum of 41 and maximum of 35,362,136. Two studies^56, 57^ had a very high number of observations, one with 35,362,136 opportunities and one with 593,118

Characteristics of all included studies are available in Supplementary material.

### Aim two: Quality of studies using observational measures

Studies with interventions intended to improve adherence to protective behaviours (n=6) were rated out of 11 relevant criteria on the NIH quality assessment checklist and studies with no interventions (n=42) were rated out of eight relevant criteria. Studies with an intervention had a median score of 10, with a range of 8-11 (Figure 1). Studies without an intervention had a median score of 7, with a range of 4-8 (Figure 2). Overall, studies in both groups generally had clearly defined study objectives, populations and variables, however very few studies reported any sample size or power estimates.

**Figure 1.**
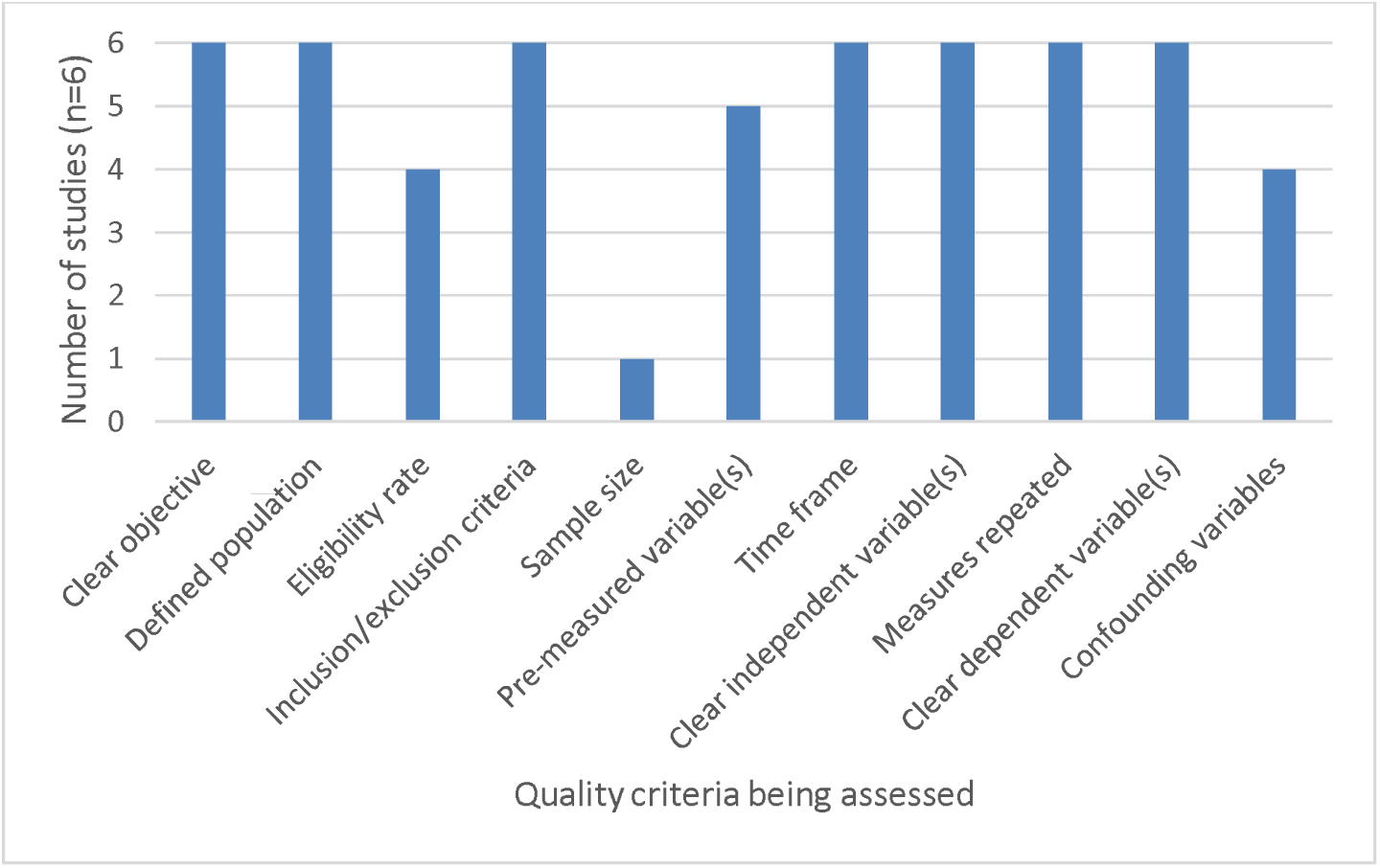
Number of intervention studies displaying relevant aspects of NIH quality assessment tool

**Figure 2.**
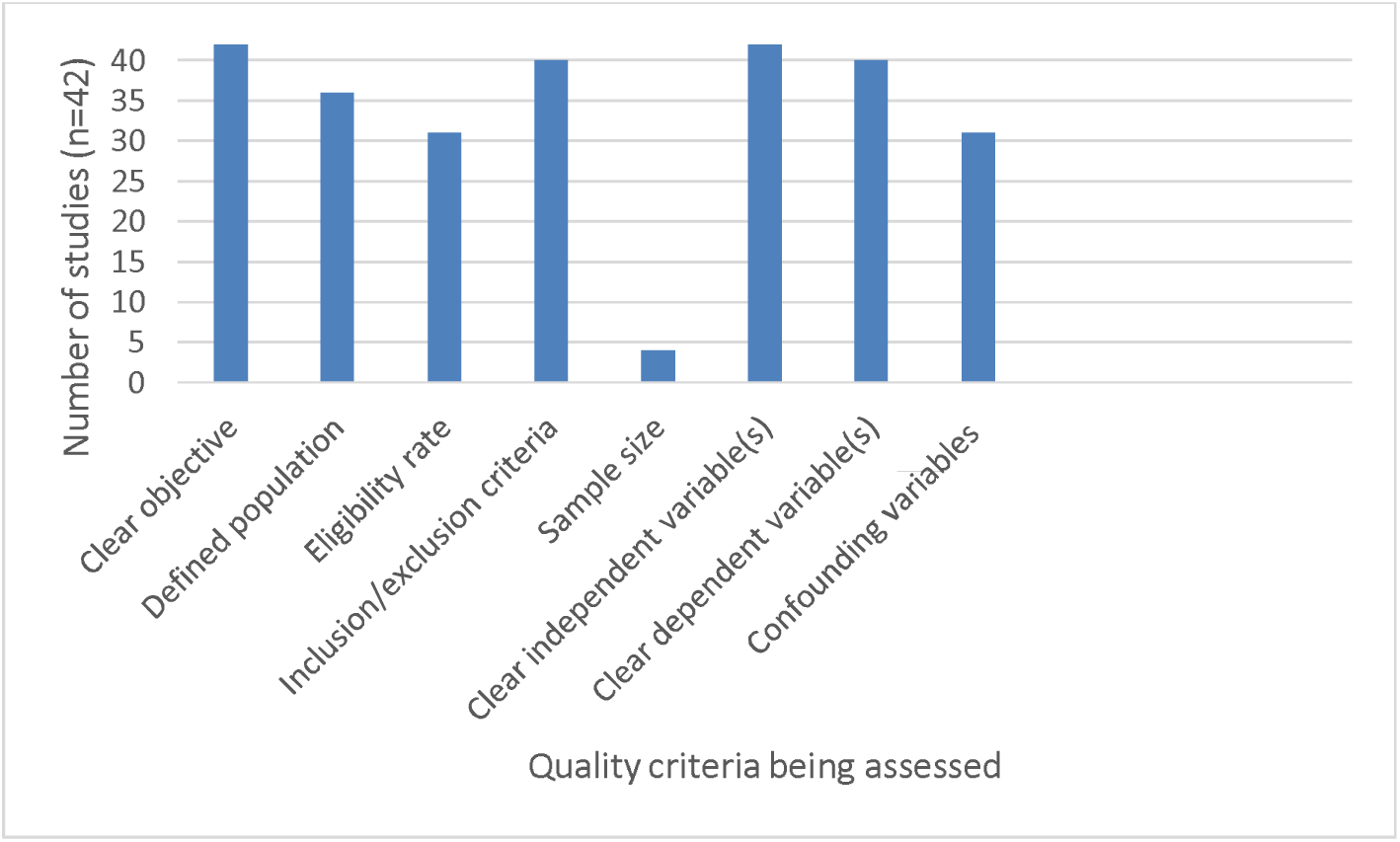
Number of non-intervention studies displaying relevant aspects of NIH quality assessment tool

### Aim three: Observational data vs self-report

In total, 21 studies contained both an observational and self-report component (see Table 1). Characteristics of all included studies are available in the Supplementary material. Three studies investigated COVID-19, while 18 investigated other infectious diseases or infectious disease practice pre-COVID-19. Of the three studies investigating behaviour during COVID-19, all three studied healthcare workers (one in Germany, one in the US and one in Thailand) and one also studied the general public (In Thailand).^26, 52, 53^. All three looked at hand hygiene adherence; one also looked at face covering use.

**Table 1.** Summary of included studies which reported an observation and self-report measure of health protective behaviour

| Study | Population; location | Observed prevalence | Self-report prevalence |
| --- | --- | --- | --- |
| Henry (1992) | ED physicians (12 staff plus rotating residents), medical students, nursing staff, and ancillary personnel; hospital emergency department | Overall face covering wearing adherence: 1.2% | Overall face covering wearing adherence: 25.5% |
| Raymond (2001) | Tattoo artists; Minneapolis and St. Paul, MN | Overall adherence was 71% | Overall adherence was 83% |
| Cohen (2002) | Dermatologists; outpatient dermatology clinics in Israel | 7 (38.5%) physicians washed hands only after contact with suspected infected material and 1 (7.7%) before each physical examination. Seven physicians (53.9%) washed their hands 1–5 times a day | 19 (37.3%) washed their hands only following contact with material suspected of being contaminated, 18 (35.3%) prior to examining every patient, 14 (27.5%) wash hands 1–5 times a day |
| Moret (2004) | Healthcare workers (physicians, nurses, and nurse assistants); French university hospital | Overall adherence was 74% | Overall adherence was 74% |
| Snow (2006) | 60 students enrolled in a certified nursing program; on day 1 and day 30 of the clinical rotation. | Day 1. Student hand hygiene:49.1%. Day 30. Student hand hygiene:52.3%. | Based on a scale of 10-100. 10 being lowest possible commitment to handwashing. On day 30 students reported 93.0 before patient contact, 95.5 after patient contact, 84.9 before donning gloves, and 95.1 after removal of gloves. |
| Alemayehu (2009) | Third and fourth year students; U.S. medical school over one academic year at the beginning of every core rotation (medicine, surgery, primary care medicine, paediatrics, obstetrics/gynaecology, neurology, psychiatry) | Overall adherence: 87.9% | Overall adherence: 87.9% |
| Soyemi, C (2010) | Doctors and nurses; three types of hospitals, i.e., public, security forces, and private, in Eastern Saudi Arabia. | Overall adherence for physicians: 27%<br>Overall adherence for nurses: 29% | Likert scale. Physicians mean self-report was 10.19/15 based on 3 hand hygiene opportunities, and nurses was 13.65/15 based on 3 opportunities. |
| Jessee (2013) | Staff on 3 medical/surgical units; a 600-bed academic medical centre (AMC) and a 110-bed community medical centre (CMC) in the South-eastern United States | Hand hygiene before glove application, was present in 14% of the CMC and 21% of the AMC staff Hand hygiene on room exit was 25% and 78% respectively. | Hand hygiene before glove application was present in 63% CMC and 50% of AMC staff. Hand hygiene on room exit 16 was 84% and 94% respectively. |
| Kim (2013) | Doctors; healthcare settings. | Adherence at baseline: 49.7%<br>Adherence in fourth quarter of 2012: 82.3% | Adherence at baseline: 82.9% Adherence in fourth quarter of 2012: 93.8% |
| Dalen (2013) | Doctors (13%), nurses (70%), housekeeping staff (9%) and visitors of patients (9%); cancer hospital. | Overall hand hygiene adherence, nurses: 47% Overall hand hygiene, doctors: 51% | Overall hand hygiene adherence, nurses: 88% Overall hand hygiene adherence, doctors: 85% |
| Lakshmi (2015) | Healthcare workers (predominantly nurses); ICU in an oncology, BMT and neurosurgical centre in South India. | Use of alcohol based hand rub 98.5%, 5 moments of hand hygiene 88.5%, 6 steps of hand hygiene 65%. | Use of alcohol based hand rub 98.5%, 5 moments of hand hygiene 88.5%, 6 steps of hand hygiene 92.5% |
| O'Donoghue (2016) | Healthcare workers, 76 radiographers, 17 nurses, and nine healthcare assistants; a radiography unit. | Overall adherence: 29 % Post-test adherence: 51% | 'I wash my hands after using the rest room' was rated a median of 5/5 on a Likert scale, quartiles 4-5 before and after intervention. |
| Galiani (2016) | General public; households and community settings in Peru | Handwashing with soap was observed in only 16% of the events that required it. 20% of faecal contact events, 25% of eating events, 6% of child feeding events, and 10% of food preparation events. | Less than 50% reported hand washing at times of faecal contact. 39% reported with toilet use, 34% cleaning up after children, 68% when cooking or with food preparation and 31% when feeding a child. |
| Keller (2018) | ED staff, 100 nurses, 13 staff emergency physicians, 25 medical interns, and various other professions (e.g. maintenance and nursing assistants). Non-ED consulting physicians and surgeons regularly visiting the ED; in an Emergency Department of the University Hospital Zurich | Hand hygiene adherence during baseline: 56%. Hand hygiene adherence during intervention: 64% | Hand hygiene adherence during baseline: 4.12. Hand hygiene during intervention 4.03 (on a Likert scale 1-5, 5 being always) |
| Baloh (2019) | Healthcare workers; 3 large academic US hospitals | Hand hygiene before gloving was performed 42% of the time | Hand hygiene before gloving was reported 88% of the time |
| Le (2019) | Healthcare workers (physicians, nurses, care assistants, and student nurses); large central hospital in Vietnam | Physicians overall hand hygiene adherence: 14.6%. Nurses overall hand hygiene adherence: 38.8% | Physicians overall hand hygiene adherence: 67.2%. Nurses overall hand hygiene adherence: 97.8% |
| Woodard (2019) | Healthcare workers (nurses, physicians, technicians) and patient interactions; 750-bed tertiary care hospital in Baltimore, Maryland. | Overall hand hygiene adherence: 35% of all opportunities. Overall entry and exit hand hygiene adherence: 90% | 81% of the sample estimated they miss performing hand hygiene when they realize it should be performed 10%-20% of the time. An additional 11% reported missing hand hygiene 30%-40% of the time. 4% estimated they missed performing hand hygiene 90%-100% of the time. |
| Kelcikova (2019) | Doctors and nurses; eight hospitals in two countries (five in Slovakia and three in the Czech Republic) | Overall adherence: 67.7% | Overall adherence: 74.0% |
| Derksen (2020) | Healthcare workers (physicians, midwives, and nurses); two German obstetric hospitals during and after the onset of the COVID-19 pandemic. | After declaration of pandemic overall hand hygiene adherence: 95%. After body fluid exposure risk: 100%. | After declaration of pandemic overall hand hygiene adherence: M = 5.03, SD = 0.75 on a 6 point scale. After body fluid exposure risk: M = 5.66, SD = 0.67. |
| Dowding (2020) | 400 U.S. home care nurses; community care organization | Overall hand hygiene adherence: 45.6%. | Overall hand hygiene adherence: 99.4% |
| Skuntaniyom (2021) | 119 healthcare workers (mostly nurses and laboratory workers) and 100 general public (mostly administrative workers and professionals); Thailand in two inpatient hospitals providing COVID-19 testing and treatment. | 100% of patients and 100% healthcare workers wore face covering correctly. 35.2% of patients and 40.0% of healthcare workers correctly cleaned all areas of both hands. | 86% of patients and 95.8% of healthcare workers wore a face covering correctly. 67% of patients and 84.9% of healthcare workers reported adhering to hand hygiene. |

Self-reported and observed hand hygiene behaviour differed, with self-reported rates being around twice that of observed rates. The biggest difference seen in hand hygiene rates was 99% self-reported and 46% observed in a study of healthcare workers engaging in community-based patient care activities in the US^52^. Observed adherence in this study varied by the activity as well as by the period during the COVID-19 pandemic when assessments were made.

Self-reported and observed face covering wearing both had high rates of adherence, with 86% self-reported adherence among patients and 95.8% among healthcare workers in one Bangkok hospital, compared to 100% adherence when observed in both groups^25^.

Of 18 studies investigating other infectious diseases, most studies (n=15) looked at hand hygiene in a healthcare worker population^63, 64, 66, 68–77, 79, 80^, while two studied it in the general public^65, 78^. One studied face covering use in healthcare workers^67^. None assessed physical distancing. Studies were conducted in Asia (n=7), North America (n=8), Europe (n=3) and South America (n=1).

Self-reported hand hygiene behaviour was higher than observed data in most studies (n=11). The greatest differences were 31% self-reported versus 6% observed hand hygiene in the general public in Peru^80^, and 67% self-reported versus 15% observed hand hygiene in healthcare workers in a large hospital in Vietnam^72^. In the only study that examined it, face covering wearing was self-reported at 25% but observed at 1% in emergency department personnel at a Minnesota public teaching hospital^67^.

In only one paper was uptake of protective behaviours lower in self-report data than observed data, and this only applied to a small subset of participants, 4% of the total sample, who expressed that their hand hygiene adherence was between 0% and 10%, compared to an observed rate of 35%. 81% of the total sample rated their hand hygiene adherence as between 80% and 90%, comparable to the 90% adherence that was observed^72^.

Self-reported rates of behaviour matched observed rates in three studies, all studying hand hygiene: one study assessing healthcare workers’ in a French university hospital^67^, one looking at medical students’ ^75^, and one looking at healthcare workers’ behaviour in an intensive care unit in South India^81^.

### Quality assessment of Non COVID-19 Studies

Studies with interventions intended to improve adherence to protective behaviours (n=2) were rated out of 11 relevant criteria on the NIH quality assessment checklist and studies with no interventions (n=16) were rated out of eight relevant criteria. Studies with an intervention had a median score of 7, with a range of 6-9 (Figure 3). Studies without an intervention had a median score of 6, with a range of 1-8 (Figure 4). Overall, studies in both groups generally had clearly defined study objectives, populations and variables, however very few studies reported any sample size or power estimates.

**Figure 3.**
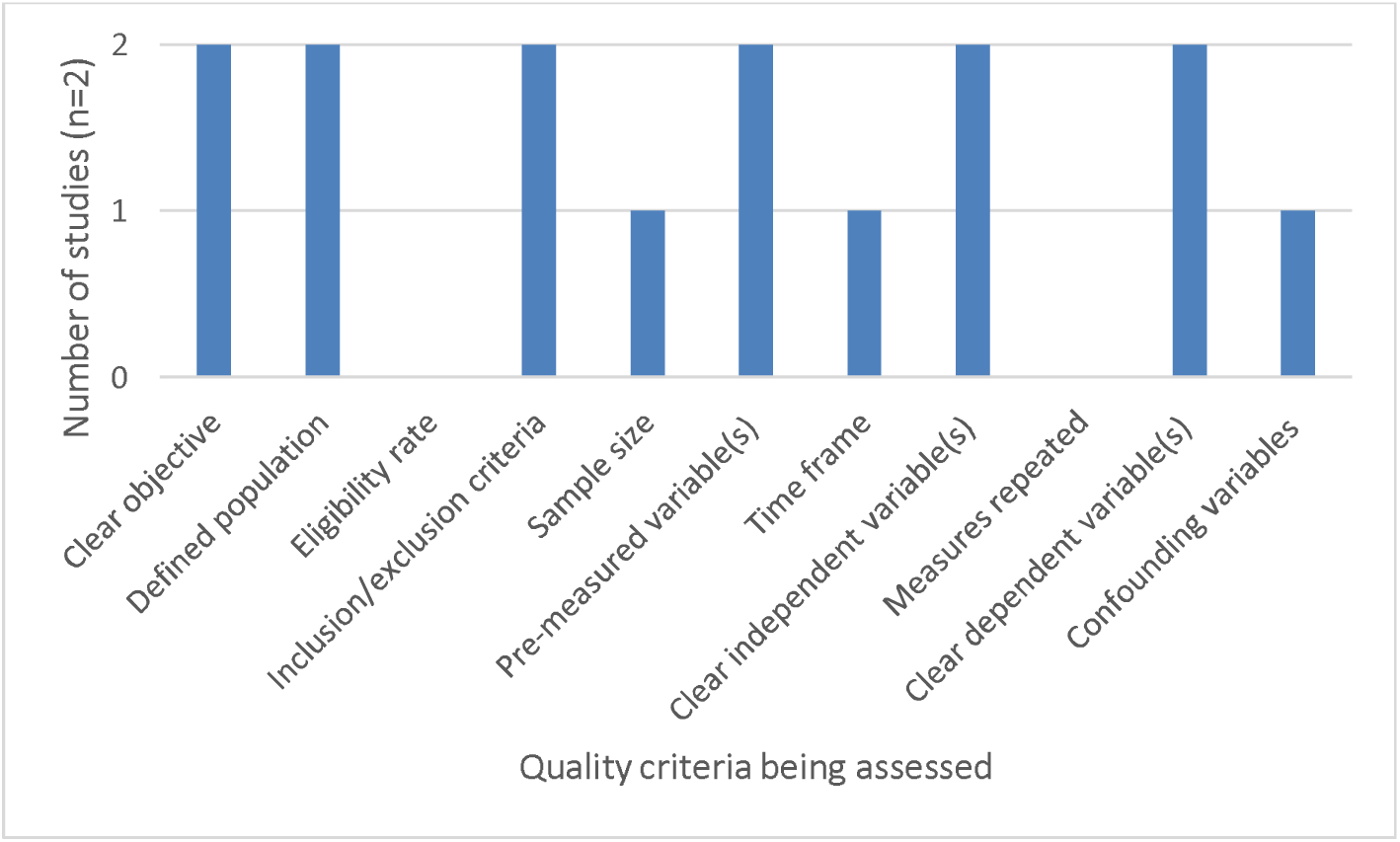
Number of non-COVID-19 intervention studies displaying relevant aspects of NIH quality assessment tool

**Figure 4.**
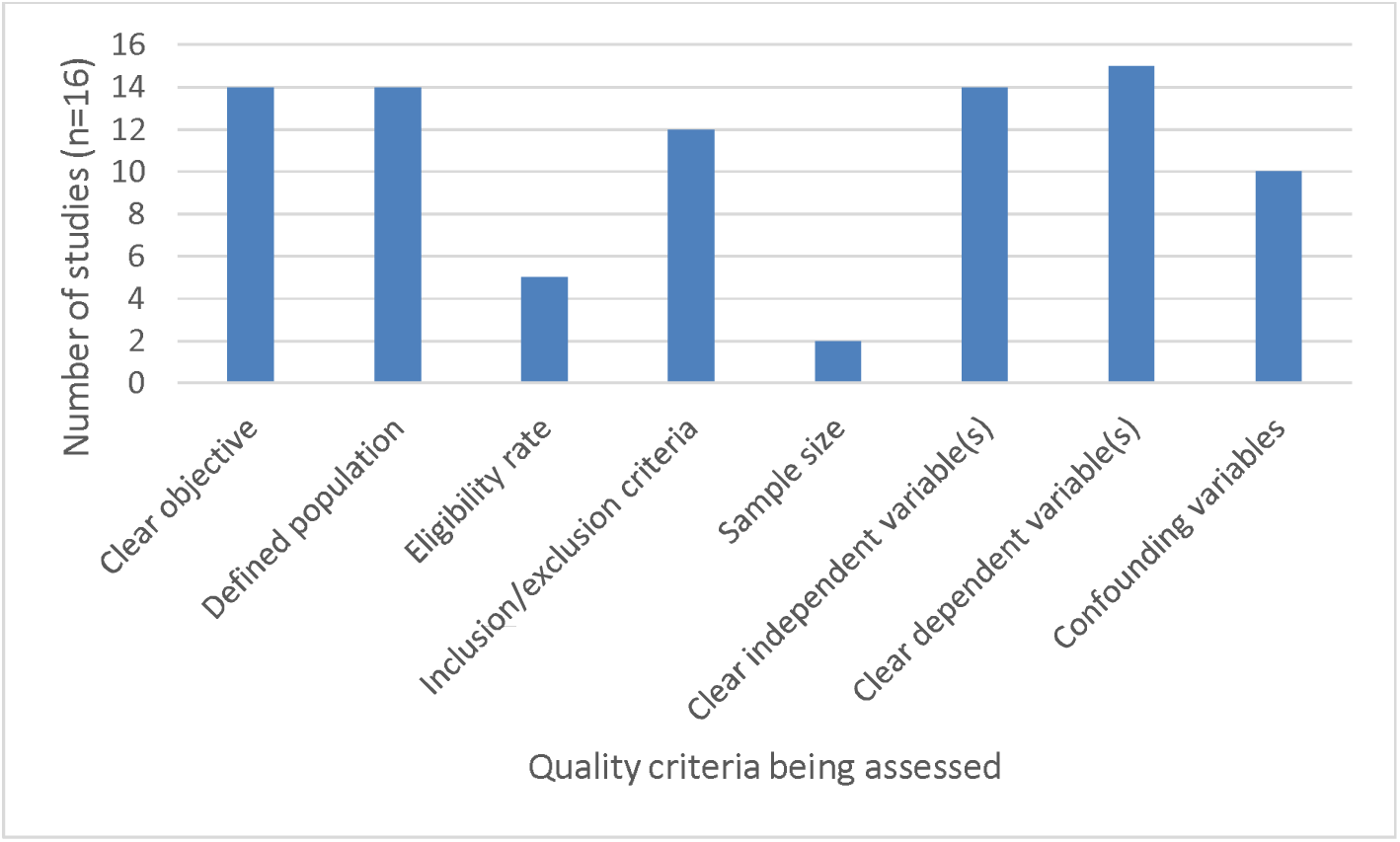
Number of non-COVID-19 non-intervention studies displaying relevant aspects of NIH quality assessment tool

## Discussion

Improving uptake of health protective behaviours is an important public health challenge, not only for COVID-19, but for infectious disease prevention more widely. Face coverings, hand hygiene and maintaining physical distance have all been identified as effective infection control strategies that have relatively few downsides in comparison to more far-reaching interventions such as society-wide ‘lockdowns.’ ^12^ Identifying ways to achieve good adherence to such measures requires that we are first able to measure adherence accurately. Though self-report can be a useful proxy for behaviour, our review suggests that academic research has become overly reliant on it, so much so that although we identified 27, 279 papers which included terms related to COVID-19 and to hand hygiene, face covering or social distancing, just 48 of these papers (<0.2%) actually studied the behaviour in question objectively.

It is likely that several factors influence the way in which behaviour is measured. Speed, cost, ease and the ability to explore associations with other variables which may be best measured via self-report (such as, for example, trust in government, exposure to conspiracy theories or the perceived efficacy of an intervention) are all valid reasons for opting to use self-report over observation. Indeed, the importance of ease as a factor likely explains why only seven studies have attempted to assess physical distancing using observational techniques while 35 have looked at face coverings: it is difficult for an observer to judge distance between two people and to identify whether people need to maintain distance from each other (e.g. if they do or do not live in the same household), but easier to assess whether they are wearing a face covering. Nonetheless, our review also points to the dangers of over-reliance on self-report. The 21 studies that we identified which compared self-report and observed behaviour repeatedly demonstrated that self-report over-estimates hand hygiene behaviour, sometimes dramatically so, while evidence for the validity of self-reported distancing and face covering use is limited in the current literature. Although outside the date range for our search, two recent pre-prints support this point for hand hygiene and extend the evidence of a self-report gap to include face covering and distancing. One study demonstrated that self-reports of “always” wearing a face covering when in specific public spaces in a national UK Government-funded survey matched observed behaviour in those locations, but that an additional 23% of people reported “sometimes” wearing face coverings in these situations, something which could not be accounted for in the observations.^81^ The other study, of a single university campus in the UK, found that while 68% of survey respondents reported always cleaning their hands while entering a university building, observation of the only entrance to the main campus building found the true rate to be 16%^82^. Reported and observed rates for distancing were also discordant (49% vs 7%) while the gap for wearing a face covering was smaller but still noticeable (90% vs 82%). While multiple factors may account for these discrepancies^83^, recall bias and social desirability would seem most likely to lead to inflated estimates of behaviours such as hand hygiene and physical distancing. The relative novelty of face coverings for many people, the limited number of occasions they need to be worn during the day for many members of the general public and their greater salience and hence ease of recall may partly mitigate these biases and explain why self-reported wearing of a covering may be a more reliable measure of behaviour than self-reported hand hygiene or distancing.

Notably, the quality of studies that included an observational measure was generally good in most respects. The one exception was that relatively few studies provided a sample size justification. We suspect this is linked to the difficulty of setting a pre-determined sample size in advance of a naturalistic study. For example, it can be difficult to predict how many people will pass by an observer over a set period of time. The relatively high quality may reflect the tendency for authors who choose to take the difficult route of evaluating behaviour via an objective measure to have also considered other ways of maximising the quality of their study.

### Suggestions for future research

Plenty of scope exists for future work to expand this literature. First, there is a pressing need to establish the validity of self-reported behaviour. At present, the limited literature that exists focusses almost entirely on hand hygiene. During the COVID-19 pandemic we found no studies comparing self-report and observational data for physical distancing and only one for the use of face coverings, although more work in this area is starting to appear^81, 82^. Future work should test approaches to improve the validity of self-report data and also test whether the correlates of self-reported behaviour (which are the basis for many policy recommendations and proposed interventions) can be replicated as correlates of observed behaviour. Consideration should be given to the potential differences in validity that may be observed across population and settings. For example, in the studies that we reviewed, observed adherence tended to be higher in studies of healthcare workers than in general population samples.

Second, our review focussed solely on three behaviours: hand hygiene, face covering use and physical distancing. While important, these are only a subset of the complex set of behaviours that members of the public have been encouraged to adopt during the COVID-19 pandemic. We have not systematically reviewed the literature on the validity of self-report measures of, for example, testing uptake or self-isolation, but have no reason to suspect that self-report is more valid for these behaviours, given that there is substantial social desirability involved and that, for some of them, research participants may technically be liable to legal action if they admit to non-adherence. Nonetheless, key studies on these outcomes rely entirely on self-report^84, 85^. The one notable exception to this list is vaccination, a memorable, binary outcome for which self-report has been shown to be reasonably, though not entirely, valid^86, 87^.

Third, while our review may give the impression that observation is a single ‘gold standard’ metric for behaviour, it is clear that there are multiple methods of observation. We identified methods including direct study of behaviours by trained observers, video observation, automated technology, and the use of newly developed technology using AI and machine learning in place of an observer. The use of such technology has been demonstrated with face covering wearing studies, as well as studies that measure crowd density with social distancing within the crowd data ^88, 89, 90^. These techniques all have their pros and cons in terms of intrusiveness, cost, capacity, ability to identify behaviours that may be partially obscured and so on. A ‘one-size-fits-all approach’ may not be possible. Nonetheless, further work to develop a set of standardised observational protocols for key outcomes may assist in promoting the use of such techniques and allowing better comparison between studies.

### Limitations

Several limitations should be considered for this systematic review. First, our conclusions are limited by the availability of data in the literature. The relative absence of observational data relating to face covering wearing or physical distancing is an important result in its own right, but also limits our ability to assess the adequacy of self-report for these behaviours. Second, while we made efforts to search widely for relevant studies, including in COVID-19 specific databases, it is possible that we missed some studies which used terminology relating to an observational method that we did not include in our search. Given the rapidity with which the COVID-19 literature has expanded, with approximately a quarter of a million papers appearing in Scopus alone in less than two years, it is likely that additional studies will have been added to the databases that we searched in the time taken between completing our search and publication of this paper.

In this review, we have not attempted to pool the rates of behaviour observed in the various studies. The differing contexts in the studies we included means that any pooled estimate would not be meaningful. For example, it is probably not useful to compare rates of observed hand hygiene among healthcare workers working on COVID-19 wards ^26^ with those among high school students attending their graduations^49^.

### Conclusions

The COVID-19 pandemic witnessed an explosion in research covering every aspect of the crisis. Within the field of behavioural science, there has been a heavy focus on ways to promote behaviours believed to reduce infection transmission. Almost all of these studies have measured whether people say they have engaged in specific behaviours. Few have measured the behaviour itself. This is problematic. For hand hygiene, observed adherence tends to be substantially lower than estimates obtained via self-report. There are few studies that have tested the validity of self-reported face covering use or physical distancing, but these also suggest that self-reports tend to be biased. Future research in this field should make greater use of observational methods where possible and should carefully consider the validity of any self-report measure where this is not possible.

## Data Availability

All data produced in the present work are contained in the manuscript

## Supplementary Material

**Figure 5:**
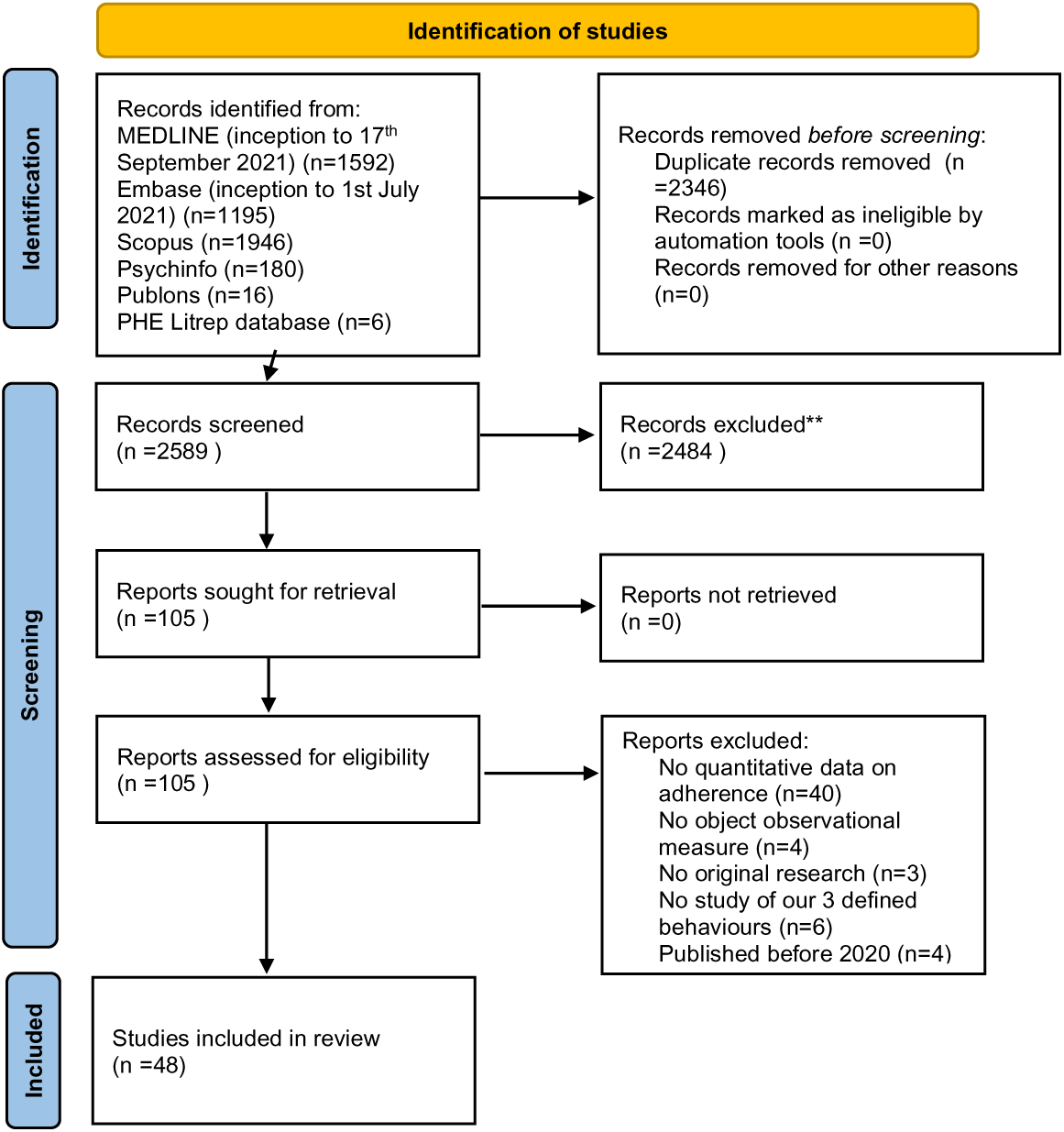
Flow chart for included studies aims one and two

**Figure 6:**
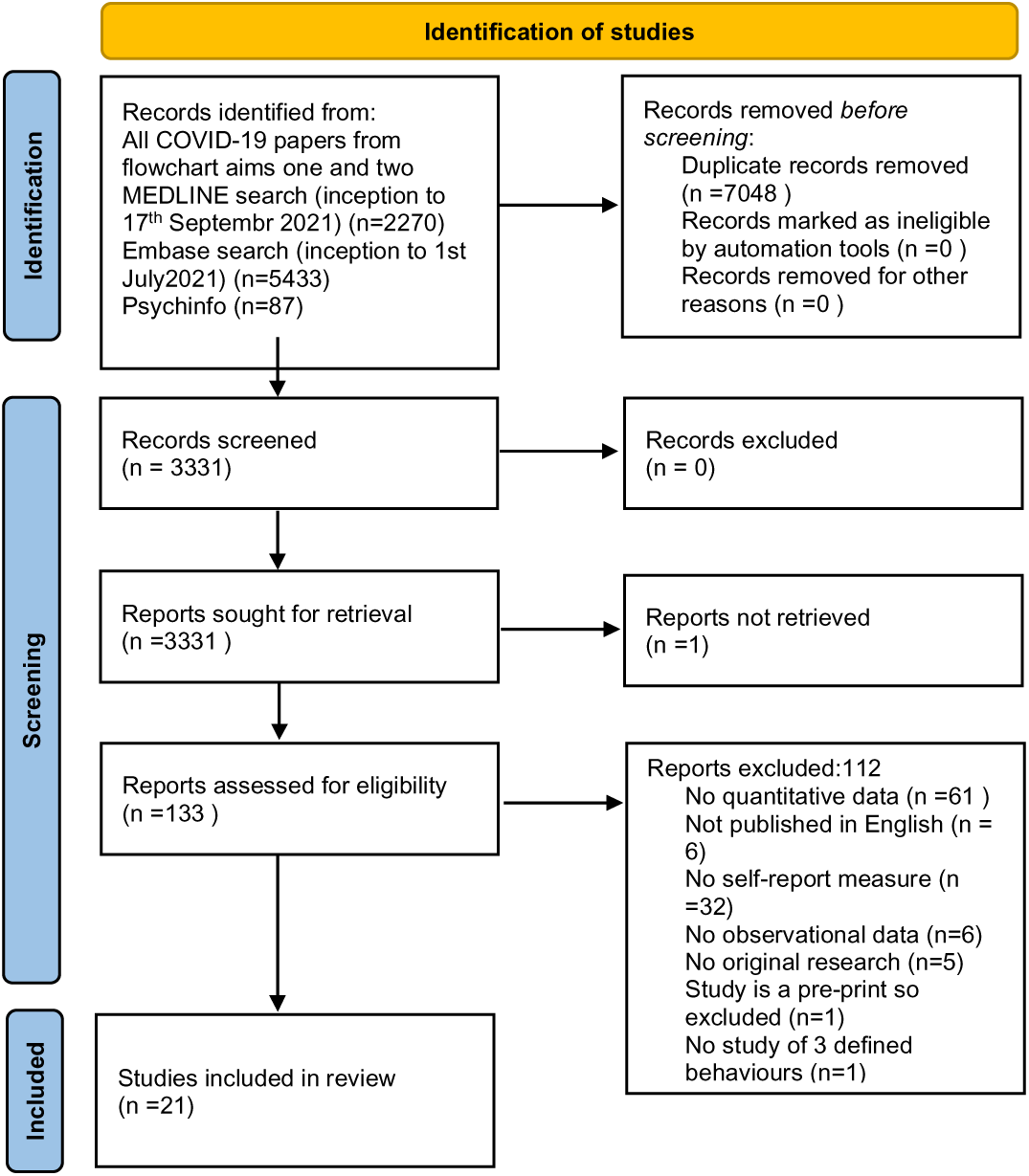
Flowchart for included studies aim three

**Table 2:** Characteristics of included studies aims one and two (COVID-19 papers)

|  | **Date of observations** | **Date of publication** | **Intervention component** | **Study design** | **Number of events** | **Sample size** | **Hand hygiene adherence %** | **Face covering adherence %** | **Social distance adherence %** | **Where did observations take place** | **Population observed** | **Other location data** |
| --- | --- | --- | --- | --- | --- | --- | --- | --- | --- | --- | --- | --- |
| 1 | 1 Jan 2015-31 Dec 2020 | Apr-21 | no | direct observer | 13494 | 13494 | 66% | n/a | n/a | hospital | healthcare workers | Sicily, Italy |
| 2 | 22 Apr-9 May 2020 | 13-Aug-20 | no | direct observer | 1000 | 1000 | n/a | 57% | n/a | public parks, banks, outpatient clinics, bus stations | general population | Iran |
| 3 | not reported | 02-Jun-20 | no | direct observer | 382 | 382 | n/a | 64.1% | n/a | hospital | healthcare workers and hospital staff | India |
| 4 | 5-8 Apr 2020 | 09-Jul-20 | no | direct observer | 78 | 78 | n/a | 32% | n/a | food stores | general population | Braga, Portugal |
| 5 | May-Jun 2020 | 18-May-21 | no | video observation | 383 | 383 | n/a | 43% | n/a | public streets | general population | Amsterdam, Netherlands |
| 6 | 5-8 Apr 20; 18-27 Apr 20; 18-27 Jun 2020 | Feb-21 | no | direct observer | 238 | 238 | n/a | 96.6% men, 100% women | n/a | food stores | general population | Braga, Portugal |
| 7 | 1-29 Feb 2020 | 25-Mar-21 | no | direct observer | 10211 | 10211 | n/a | 82.5% | n/a | public streets | general population | Hong Kong |
| 8 | 10, 18, 25 May 2020 | 23-Feb-21 | no | direct observer | 3965 | 2353 | n/a | 73.6% at time 1; 66.5% at time 2; | n/a | public streets, parks, enclosed | general population | Poland |
|  |  |  |  |  |  |  |  | 65.7% at time 3; highest while shopping (84.9%; 81.7 and 80.3%) and the lowest during outdoor sport activity (63%; 53.5 and 48.7%). |  | shopping centres |  |  |
| 9 | Sep-Nov 2020 | 05-Feb-21 | no | direct observer | 17200 | 17200 | n/a | 76.70% | n/a | universities | staff and students | rural and urban universities across USA |
| 10 | 20 Jul 2020 for 4 weeks; 21 Sep 2020 for 14 weeks | 03-May-21 | yes | direct observer | 4122 | 4122 | n/a | 82.2% pre intervention; 92.2% wore after intervention | n/a | hospitals | healthcare workers | Connecticut, USA |
| 11 | Jul-Aug 2020; 23-30 Nov 2020; 1-5 Dec 2020 | 18-Apr-21 | yes | direct observer | 219 | 219 | 35.2%; 40% healthcare workers | 100% of public and healthcare workers | n/a | hospitals | patients and healthcare workers | Bangkok, Thailand |
| 12 | May-20 | 01-Sep-20 | no | direct observer | 271 | 271 | n/a | 94.2% overall. 100% on COVID-19-designated wards and<br>48% on<br>non-COVID-19 wards | n/a | hospital | healthcare workers | UK |
| 13 | 3-9 Jun<br>2020; 24<br>Jul-3 Aug<br>2020 | 15-Oct-20 | no | direct<br>observer | 9935 | 9935 | n/a | 41.5% | n/a | retail<br>stores | general<br>population | rural and<br>urban<br>Wisconsin,<br>USA |
| 14 | 16-30<br>May 2020 | 13-Apr-21 | no | direct<br>observer | 1004 | 1004 | n/a | 75.5% | n/a | public<br>business<br>es | general<br>population | Vermont,<br>USA |
| 15 | 5-8 May<br>2020 | 03-Aug-20 | no | direct<br>observer | 892 face<br>cover<br>wearing, 871<br>social<br>distancing | 1641 | n/a | 12.6% | 98%<br>adherence | vehicles | drivers and<br>passengers | Ghana |
| 16 | 30 Mar-12<br>Apr 2020 | Sep-20 | no | direct<br>observer | 3322 | 3322 | n/a | 98.2%<br>wore face<br>cover;<br>92.3%<br>wore face<br>cover<br>correctly | n/a | market,<br>hospital | general<br>population | Malaysia |
| 17 | 12 days<br>9am-8pm<br>during<br>'phase 2'<br>in Italy | 01-Jan-21 | no | direct<br>observer | 1036 | 1036 | n/a | 25.5% | n/a | in the<br>street',<br>most<br>people<br>observe<br>d were<br>outside<br>shops,<br>waiting<br>to enter | general<br>population<br>(excluded<br>children and<br>'people with<br>special<br>needs' | Italy |
| 18 | 11am to<br>1pm 6-9<br>Aug 2020 | 17-Nov-20 | yes | video<br>observati<br>on quasi<br>experime<br>nt; | 1286 | 400 | n/a | n/a | 20% | a universit<br>y canteen | university<br>canteen<br>customers | Thailand |
|  |  |  |  | comparative<br>behavioural<br>observation |  |  |  |  |  |  |  |  |
| 19 | 4-25 May 2020 | 03-May-21 | no | direct observer | 182 | 182 | n/a | 94% | n/a | on public transport (subway and local trains) | public transportation users | Paris, France |
| 20 | 21 Sep 2020 to 2 Oct 2020 | 24-Oct-20 | no | direct observer | 187 | 187 | n/a | 80.1% at perimeter, 93.4% by entry | n/a | general ob/gyn outpatient clinic | patients and visitors | North Carolina, USA |
| 21 | 15-18 Jun 2020; 20-21 Jun 2020; 22-24 Jun 2020 | 14-Oct-20 | yes | direct observer-quasi experiment; comparative behavioural observation | 466 | 466 | 0% adherence pre intervention; 12% adherence post intervention | n/a | n/a | hospital | outpatients and visitors | Thailand |
| 22 | 2-11 Aug 2020 | 14-Jan-21 | no | direct observer | 10440 | 10440 | n/a | 45.6% wore face cover; 75.6% of face covering wearers wore it correctly | n/a | in the street | general population | 8 urban districts, 92 neighbourhoods of Ahvaz, Iran |
| 23 | Jan 2018-October | 29-Jul-20 | no | video observation | 7586 | 7586 | n/a | face cover wearing | n/a | public transport | general population | China, Japan, |
|  | 2019;<br>Feb-Mar<br>2020 |  |  | on |  |  |  | increased<br>in all<br>regions<br>except the<br>US, from<br>(1.1%) to<br>(99.4%) in<br>mainland<br>China.(3.1<br>%) to<br>(38.7%) in<br>Japan<br>(0.8%) to<br>(85.5% )<br>in South<br>Korea.<br>(0.2%) to<br>(1.6%) in<br>Western<br>Europe<br>and<br>(0.4%) to<br>(2.1%) in<br>the US |  | ation<br>stations,<br>streets,<br>parks |  | South<br>Korea, US,<br>England,<br>France,<br>Germany,<br>Spain, Italy |
| 24 | 5-7 Mar<br>2020 | 06-Jun-20 | no | direct<br>observer | 1761 | n/a | 79.44% | n/a | n/a | hospital | healthcare<br>workers | Tongji,<br>China |
| 25 | 27 Feb-21<br>Apr 2020 | 10-Dec-20 | no | direct<br>observer | 610 | 127 | 82%<br>adhere<br>nce in<br>COVI<br>D-19ward<br>vs 65%<br>non-COVI<br>D-19<br>ward | n/a | n/a | hospital<br>pulmono<br>logy<br>departm<br>ent | healthcare<br>workers | Cologne,<br>Germany |
| 26 | 21-30 Apr<br>2020 | 26-Oct-20 | no | direct<br>observer | 468 | 468 | n/a | 85.42%<br>wore face covering;<br>66% wore<br>face<br>covering<br>correctly; | n/a | street,<br>public transport<br>ation,<br>grocery<br>store<br>line | general<br>population | Basseterre,<br>St. Kitts |
| 27 | Jul-Aug<br>2020 | 12-Mar-21 | no | direct<br>observer | 422 | 422 | n/a | 54.68% | n/a | taxi<br>stands | taxi drivers | Dessie City<br>and<br>Kombolcha<br>Towns,<br>Ethiopia |
| 28 | 20-24 Jul<br>2020 | 28-Dec-20 | no | direct<br>observer | 2302 | 2286 | n/a | 98% | n/a | ophthal<br>mology<br>outpatie<br>nt<br>clinics | patients | Pennsylvan<br>ia, USA |
| 29 | Apr-Jun<br>2020 | 03-May-21 | no | direct<br>observer | 2080 | 2080 | n/a | 65% | n/a | outside<br>grocery<br>stores | general<br>population | Western<br>Australia |
| 30 | Jul-20 | 05-Jan-21 | no | direct<br>observer | 1096 | 1096 | n/a | 70.2% at<br>mosque;<br>68.7% in<br>malls;<br>32.6% in<br>park;<br>45.2% in<br>barbersho<br>p | n/a | in the<br>commun<br>ity<br>(mosque<br>s, malls,<br>parks,<br>barbersh<br>ops) | general<br>population | cities in<br>Jazan<br>region of<br>Saudi<br>Arabia |
| 31 | 30-Jul-20 | 01-Sep-20 | no | direct<br>observer | 200 | 100 | n/a | 77% | n/a | outdoor<br>street<br>location<br>s | general<br>population | Waikiki and<br>Honolulu,<br>Hawaii,<br>USA |
| 32 | Jun-20 | 18-Nov-20 | no | direct<br>observer | 431 | 431 | 57.46%<br>adhere<br>nce on<br>subway<br>44.78%<br>adhere<br>nce<br>when shopp<br>ng | 78.35% on<br>subway;<br>46.01%<br>when<br>shopping | n/a | subway<br>station<br>and a<br>shop | general<br>population | Tehran, Iran |
| 33 | 25 Jun-21<br>Jul 2020 | 29-Mar-21 | no | direct<br>observer | 3354 | 3354 | n/a | 56.4%<br>wore face<br>coverings;<br>75.2% of<br>face<br>coverings<br>were worn<br>correctly | n/a | inside<br>shoppin<br>g<br>centres<br>and train<br>stations;<br>outside<br>shoppin<br>g<br>centres | general<br>population | 13 cities in<br>Pays de la<br>Loire,<br>France |
| 34 | 4 May-11<br>Jun 2020 | 25-Jan-21 | no | direct<br>observer | 3061 | 3061 | 2% | 96%<br>smokers<br>possessed<br>a face<br>covering.<br>While<br>smoking,<br>0.2% wore<br>there face<br>covering<br>fully,<br>81.6% of<br>smokers<br>put the<br>face cover<br>under the<br>chin and<br>13.8%<br>carried it<br>in the<br>hand,<br>32.4% did<br>not wear a<br>face<br>covering<br>immediate<br>ly after smoking. | 25.7% | outdoor<br>street<br>location<br>s | general<br>population | Hong Kong |
| 35 | Jul-20 | 24-Jan-21 | no | mixed methods. direct observer/ media | 1152 | 1152 people | n/a | 70% | 0 | in person high school graduation | high school students | USA |
| 36 | unstated, during COVID-19 | Dec-20 | no | video observation | 1594 | 1594 | n/a | n/a | 40.4% Oxford Town Centre dataset; 17.6% in the Mall dataset; and, 58.1% CUHK Square dataset | oxford town centre | all general public any age | city centre/shopping mall UK |
| 37 | 2 January to 15 March 2020 | Oct-21 | no | direct observer | 267 | unknown | 47 per cent pre-COVID-19 pandemic to 95 per cent just before lockdown | n/a | n/a | obstetric ward | healthcare worker | Germany |
| 38 | unknown | Dec-20 | no | direct observer and survey | 400 | 400 people | 45.6% | n/a | n/a | home health care | nurses | USA |
| 39 | June 1, 2020, to | Oct-20 | yes | direct observer | 42 | 42 people | 33.59% | n/a | n/a | tertiary care | surgical trainees | Calcutta |
|  | July 1, 2020 |  |  |  |  |  |  |  |  | hospital |  |  |
| 40 | unknown | Jan-21 | no | direct observer | 1053 | unknown | 15% | 68.2% | n/a | private and government-subsidized RCHs | healthcare worker | Hong Kong |
| 41 | February 7 to 25, 2020 | 2020 | no | direct observer | 11680 | 12208 people | n/a | for women, 70.5% and men 50% | n/a | residential areas, shopping districts, office quarters | adults and children over 2 in general population. | Taipei and major satellite cities in New Taipei |
| 42 | 10 weeks before the time of the school closures (the week beginning January 5 through the week beginning March 8) with the 10-week period after school closures (the week beginning March 15 through the week beginning<br>May 17). | Jan-21 | no | automated technology hand sanitizer event measurement | 35,362,136 | unknown | 48.52% before the school closures to 58.05% after | n/a | n/a | 9 US hospitals | healthcare workers/general public | USA |
| 43 | Mar 2020<br>- Jun<br>2020) | Nov-20 | no | mixed<br>methods.<br>automate<br>d<br>technolog<br>y hand<br>sanitizer<br>event<br>measurem<br>ent and<br>observer | 593,118 | unknown | 85%<br>hospita<br>l wide<br>and<br>90%<br>on<br>COVI<br>D-19<br>wards | n/a | n/a | inpatient<br>areas of<br>the<br>hospital | healthcare<br>workers | USA |
| 44 | April to July 2020 | Apr-21 | yes | direct observer | 60 | 60 | n/a | n/a | 78.3% | regional hospital in Korea. | healthcare workers | Korea |
| 45 | March and April 2020 | April 2020 | no | direct observer | unknown | unknown | 97% | n/a | n/a | tertiary care hospital Pakistan | healthcare workers | Pakistan |
| 46 | Oct-20 | Jul-20 | no | direct<br>observer | 300 | 300 | 85% | 74% | 86% | paediatric<br>tertiary<br>care<br>centre | parents of<br>children on<br>arrival to<br>hospital | India |
| 47 | unknown | Sep-20 | no | direct observer | 400 | 48 | 45.6%. highest after contact with body fluid (65.1% ) and lowest after touching a patient (29.5% ). | n/a | n/a | home health care | healthcare workers | USA |
| 48 | Jul-20 | Feb-21 | no | video<br>recording | 780 | 780 | n/a | 61% | 69% | Arches<br>National<br>Park | general<br>public | USA |

**Table 3:** Characteristics of included studies aim 3 (Non COVID-19)

| **Study** | **Date of observation** | **Date of pub** | **Study design** | **Number of events** | **Sample size** | **Hand hygiene adherence observed** | **Self-reported prevalence** | **Where did observations take place** | **Population observed** | **Other location data** |
| --- | --- | --- | --- | --- | --- | --- | --- | --- | --- | --- |
| 1 | unknown | 2006 | direct observer/survey | 890 | 60 | period 1, student hand hygiene was 49.1%, mentors 44.3 % during observational period 2, student hand hygiene was 52.3%, mentor 53.3%. | students reported 92.98 before patient contact, 95.53 for commitment to handwashing after patient contact, 84.89 before donning gloves, and 95.11 after removal of gloves. Based on a scale of 10-100. 10 being lowest possible. | hospital setting | sixty students enrolled in a certified nursing program were selected to participate in the study. | USA |
| 2 | unknown but questionnaire was 1 year before observations | 2002 | questionnaire, observation and hand bacteria measurement | 555 observed hand hygiene opportunities | 13 observed, 51 survey responses (from a conference - not the same people) | 31.4% of all opportunities; 53.9% 1-5 times a day; 38.5% only after contact with infected material; | 37.3% following contact with contaminant; 35.3% prior to every patient examination; 27.5% 1-5 times a day 3.9% following every | two outpatient dermatology clinics | physicians | Israel |
|  |  |  |  |  |  | 7.7% before each physical examination | patient exam; 3.9% once per hour |  |  |  |
| 3 | October 1999-April 2000 | 2001 | questionnaire, observation | not reported | 61 artists surveyed, 25 of whom were observed | 71% | 83% | 29 professional tattoo studios | professional tattooists | the seven-county metropolitan area of Minneapolis and St. Paul, Minnesota |
| 4 | Sep-02 | Nov-04 | questionnaire, observation | not reported | 1050 survey responses, 206 observed | 74% | 74% | 25 care units within a University Hospital | medical staff (physicians, nurses, nursing assistants) | Nantes University Hospital, France |
| 5 | July-Sept 1989 | Jan-92 | questionnaire, observation | 1,018 procedures were reported x 4 outcomes = 4072 | 66 survey responses, 88 staff working (not reported how many were observed) | face covering 1.2%; | face coverings: 25.5%; | emergency department of a university hospital | medical staff (12 staff physicians, five to ten resident physicians, physician assistants registered nurses, nursing assistants and other ancillary and support staff) healthcare workers | urban, the Minneapolis/StPaul (twin cities) metropolitan area |
| 6 | 2016 observations, 6 months later (2017) interviews | 2019 | interview and observation | 4957 hand hygiene opportunities | 25 interviews, observed not stated | hand hygiene before gloving was performed 42% | 88% | 3 teaching hospitals, including 33 units (haemodialysis units, paediatric wards, intensive care units (ICUs), and emergency departments (EDs)) | (physician, physician assistant, nurse, nursing assistant, nurse practitioner) | eastern and midwestern USA |
| 7 | Nov-13 | March-May | mixed methods, self- |  | 51 | hand hygiene | hand hygiene before glove | 3 medical/surgic | registered nurses | USA |
|  |  | 2012 | report and direct observed by observer | 91 |  | before glove application, was present in 14% of the CMCs and 21% of the MDAMCs | application CMCs (63%) MDAMCs (50%). hand hygiene on room exit CMCs (84%) MDAMCs (94%) | al units at 2 medical centres within the south-eastern United States. One was a 44-bed medical/surgical unit at a 110-bed community medical centre (CMC) and the other 2 units were 28-bed medical/surgical units at a 600 bed magnet-designated academic medical centre (MDAMC) | comprised 78% of the total sample of respondents. Although physicians and therapy providers were invited to participate, none completed the survey. Therefore, results focus on licensed and unlicensed nursing staff. |  |
| 8 | unknown | Feb-19 | mixed methods: direct observer and survey | 997 | 371 survey participants. 997 people observed | physicians had the lowest rate of compliance at 14.6%, while nurses had the highest rate of compliance at 38.8%. | physicians had the lowest rate (67.2%) of self-reported hand hygiene compliance, while nurses had the highest rate (97.8%), | 11 inpatient service departments were chosen for the study: Five internal medicine services, five paediatrics services, and one intensive care unit service | the subjects of the study were healthcare workers of different positions: physicians, nurses, care assistants, and student nurses. | Vietnam |
| 9 | April and June 2016 | May 2019 | direct observers and | 811 opportunities | 87 | overall hand hygiene | self-reported hand hygiene | University Hospital | healthcare workers | Switzerland |
|  |  |  | survey | observed. A total of 87 questionnaires were returned |  | compliance was 56% during baseline and 64% during intervention period, respectively. | compliance did not differ between baseline and intervention (4.12 vs 4.03) on a Likert scale 1-5, 5 being always | Zurich |  |  |
| 10 | July-August 2016 | May-19 | mixed methods: direct observer and survey, interviews | 302. | 104 observed, 218 survey | hand hygiene was 35% of all opportunities. Overall entry and exit hand compliance was 90% | 81% of the sample estimating they miss performing hand hygiene when they realize it should be performed 10%-20% of the time. An additional 11% reported missing hand hygiene 30%-40% of the time. 4% estimated they missed performing hand hygiene 90%-100% of the time. | ICU at the University of Maryland Medical Center, a 750-bed tertiary care hospital in Baltimore, Maryland. | healthcare workers | USA |
| 11 | unknown | Oct-18 | mixed methods, self-report and direct observed by observer | 249 observed hand hygiene | 87 participants (46 medical doctors, 21 from the public hospital, 9 from the | physicians (27%) and nurses (29%) | Likert scale. Physicians mean self-report was 10.19/15 based on 3 hand hygiene opportunities, and nurses was | outpatient examination rooms and emergency departments of three types of hospitals | 87 physicians and nurses recruited while on duty during the scheduled observation periods, with each healthcare | Eastern region of Saudi Arabia |
|  |  |  |  |  | private clinic, and 16 from the Security Forces; and 41 nurses, 14 from the public hospital, 9 from the private clinic, and 18 from the security forces hospital) were recruited. Of the 87 participants who completed the questionnaire , 83 participants were observed during exams with at least three patients and hand hygiene practices were recorded using the checklist |  | 13.65/15 based on 3 opportunities. |  | worker being observed during individual medical examinations with at least three patients |  |
| 12 | unknown | 2011 | survey self-report vs | unknown | 160 medical students in | direct observations | 87.90% | hospital setting | medical students | USA |
|  |  |  | direct observation |  | year 3 and 98 in year 4 | reported proper hand hygiene in 87.9% |  |  |  |  |
| 13 | fourth quarter of 2011 to 82.3% in fourth quarter of 2012 | Jun-13 | mixed methods, self-report and direct observed by observer | unknown | unknown | the overall hand hygiene compliance rate increased from a baseline of 49.7% in fourth quarter of 2011 to 82.3% in fourth quarter of 2012 | increased from a baseline of 82.9% to 93.8% | healthcare settings | healthcare workers doctors | Korea |
| 14 | Aug-12 | 2013 | mixed methods, self-report and direct observed by observer | 470 minutes of observations in total | the observed population was 13% doctors, 70% nurses, 9% housekeeping staff, 9% visitors of patients. Questionnaires were 35 nurses and 34 doctors. | nurses: 47% actual observed compliance with hand hygiene, doctors: 51% actual observed compliance | nurses: 88% compliance with hand hygiene, doctors: 85% compliance | modern oncology hospital | medical staff (nurses, doctors) and some housekeeping staff and visitors | India |
| 15 | unknown | 2016 | direct observer/survey | 457 | 102 | 29% overall compliance. post-test compliance increased to 51 % | "i wash my hands after using the rest room" was rated a median of 5/5 on a Likert scale, quartiles 4-5 before and after intervention | hospital setting | (healthcare workers, comprising 76 radiographers, 17 nurses, and nine healthcare assistants (HCA), agreed to participate in the study. | Hong Kong |
| 16 | the baseline survey was held from May through August 2008 and covered a total of 3576 households. The follow-up survey was conducted following the completion of the project activities (March–June 2011). | Oct-15 | survey self-report vs direct observation | unknown | 3576 in survey, 45-60 households observed | handwashing with soap was observed in only 16% of the events that required it. 20% of faecal contact events, 25% of eating events, 6% of child feeding events, and 10% of food preparation events. | although almost all caregivers reported having washed their hands with soap at least once during the previous 24 h, fewer than half confirmed having carried out so at times of faecal contact (39% of caregivers associated handwashing with soap with toilet use and 34% with cleaning up after children). cooking or food preparation (68%), but lower when feeding a child (31%). | households | general public | Peru |
| 17 | unknown | Jun-15 | survey self-report vs direct observation | unknown | sixty nine respondents participated in the survey which predominantly included the nurses | use of alcohol based hand rub 94.2%, five moments of hand hygiene 88.5%, 6 | use of alcohol based hand rub 98.5, five moments of hand hygiene Q 88.5, 6 steps of hand hygiene Q-92.5 | in ICU in an oncology, BMT and neurosurgical centre in South India | healthcare workers mostly nurses | South India |
|  |  |  |  |  |  | steps of hand hygiene 65%. |  |  |  |  |
| 18 | 2015-2016 | Feb-2019 | survey self-report vs direct observation | 127 | 639,127 of these healthcare workers had been also directly observed. | 67.7% | 74% | hospital setting | healthcare workers physicians and nurses | 5 hospitals in Slovakia and 3 in Czech republic |

**Search Strategy**

EMBASE SEARCH COVID-19 papers Aim 1 and 2

1. exp coronavirus disease 2019/
2. exp Severe acute respiratory syndrome coronavirus 2/
3. exp Coronavirinae/
4. COVID-19-19.mp. [mp=title, abstract, heading word, drug trade name, original title, device manufacturer, drug manufacturer, device trade name, keyword, floating subheading word, candidate term word]
5. coronavirus.mp. [mp=title, abstract, heading word, drug trade name, original title, device manufacturer, drug manufacturer, device trade name, keyword, floating subheading word, candidate term word]
6. COVID-19.mp. [mp=title, abstract, heading word, drug trade name, original title, device manufacturer, drug manufacturer, device trade name, keyword, floating subheading word, candidate term word]
7. 1 or 2 or 3 or 4 or 5 or 6
8. exp hand washing/
9. (hand adj3 wash*).mp. [mp=title, abstract, heading word, drug trade name, original title, device manufacturer, drug manufacturer, device trade name, keyword, floating subheading word, candidate term word]
10. (hand adj3 hygiene).mp. [mp=title, abstract, heading word, drug trade name, original title, device manufacturer, drug manufacturer, device trade name, keyword, floating subheading word, candidate term word]
11. (hand adj3 sanit*).mp. [mp=title, abstract, heading word, drug trade name, original title, device manufacturer, drug manufacturer, device trade name, keyword, floating subheading word, candidate term word]
12. (hand adj3 disinect*).mp. [mp=title, abstract, heading word, drug trade name, original title, device manufacturer, drug manufacturer, device trade name, keyword, floating subheading word, candidate term word]
13. hand-sanit*.mp. [mp=title, abstract, heading word, drug trade name, original title, device manufacturer, drug manufacturer, device trade name, keyword, floating subheading word, candidate term word]
14. (hand adj3 clean*).mp. [mp=title, abstract, heading word, drug trade name, original title, device manufacturer, drug manufacturer, device trade name, keyword, floating subheading word, candidate term word]
15. hand-wash*.mp. [mp=title, abstract, heading word, drug trade name, original title, device manufacturer, drug manufacturer, device trade name, keyword, floating subheading word, candidate term word]
16. hand-hygiene.mp. [mp=title, abstract, heading word, drug trade name, original title, device manufacturer, drug manufacturer, device trade name, keyword, floating subheading word, candidate term word]
17. 8 or 9 or 10 or 11 or 12 or 13 or 14 or 15 or 16
18. exp face mask ventilation/ or exp mask/ or exp cloth mask/ or exp face mask/ or exp pediatric face mask/ or exp surgical mask/
19. (face adj3 cover*).mp. [mp=title, abstract, heading word, drug trade name, original title, device manufacturer, drug manufacturer, device trade name, keyword, floating subheading word, candidate term word]
20. mask*.mp.
21. (wear adj3 mask).mp. [mp=title, abstract, heading word, drug trade name, original title, device manufacturer, drug manufacturer, device trade name, keyword, floating subheading word, candidate term word]
22. (facial adj3 cover*).mp. [mp=title, abstract, heading word, drug trade name, original title, device manufacturer, drug manufacturer, device trade name, keyword, floating subheading word, candidate term word]
23. (facial adj3 mask).mp. [mp=title, abstract, heading word, drug trade name, original title, device manufacturer, drug manufacturer, device trade name, keyword, floating subheading word, candidate term word]
24. facial-cover*.mp. [mp=title, abstract, heading word, drug trade name, original title, device manufacturer, drug manufacturer, device trade name, keyword, floating subheading word, candidate term word]
25. (wore adj3 mask).mp. [mp=title, abstract, heading word, drug trade name, original title, device manufacturer, drug manufacturer, device trade name, keyword, floating subheading word, candidate term word]
26. 18 or 19 or 20 or 21 or 22 or 23 or 24 or 25
27. exp social distancing/
28. (social* adj3 distanc*).mp. [mp=title, abstract, heading word, drug trade name, original title, device manufacturer, drug manufacturer, device trade name, keyword, floating subheading word, candidate term word]
29. (physical* adj3 distanc*).mp. [mp=title, abstract, heading word, drug trade name, original title, device manufacturer, drug manufacturer, device trade name, keyword, floating subheading word, candidate term word]
30. (safe adj3 distanc*).mp. [mp=title, abstract, heading word, drug trade name, original title, device manufacturer, drug manufacturer, device trade name, keyword, floating subheading word, candidate term word]
31. 27 or 28 or 29 or 30
32. CCTV.mp.
33. (closed adj3 circuit adj3 television).mp. [mp=title, abstract, heading word, drug trade name, original title, device manufacturer, drug manufacturer, device trade name, keyword, floating subheading word, candidate term word]
34. exp videorecording/
35. (surveillance adj3 camera).mp. [mp=title, abstract, heading word, drug trade name, original title, device manufacturer, drug manufacturer, device trade name, keyword, floating subheading word, candidate term word]
36. 32 or 33 or 34 or 35
37. exp observational study/ or exp observational method/
38. observ*.mp. [mp=title, abstract, heading word, drug trade name, original title, device manufacturer, drug manufacturer, device trade name, keyword, floating subheading word, candidate term word]
39. 37 or 38
40. 17 or 26 or 31
41. 7 and 40
42. 39 and 41
43. 7 and 36 and 40
44. 42 or 43

MEDLINE SEARCH COVID-19 papers Aim 1 and 2

1. exp COVID-19-19/
2. exp SARS-CoV-2/
3. exp Coronavirus/
4. COVID-19-19.mp. [mp=title, abstract, original title, name of substance word, subject heading word, floating sub-heading word, keyword heading word, organism supplementary concept word, protocol supplementary concept word, rare disease supplementary concept word, unique identifier, synonyms]
5. coronavirus.mp. [mp=title, abstract, original title, name of substance word, subject heading word, floating sub-heading word, keyword heading word, organism supplementary concept word, protocol supplementary concept word, rare disease supplementary concept word, unique identifier, synonyms]
6. COVID-19.mp. [mp=title, abstract, original title, name of substance word, subject heading word, floating sub-heading word, keyword heading word, organism supplementary concept word, protocol supplementary concept word, rare disease supplementary concept word, unique identifier, synonyms]
7. 1 or 2 or 3 or 4 or 5 or 6
8. exp Hand Disinfection/
9. (hand adj3 wash*).mp. [mp=title, abstract, original title, name of substance word, subject heading word, floating sub-heading word, keyword heading word, organism supplementary concept word, protocol supplementary concept word, rare disease supplementary concept word, unique identifier, synonyms]
10. (hand adj3 hygiene).mp. [mp=title, abstract, original title, name of substance word, subject heading word, floating sub-heading word, keyword heading word, organism supplementary concept word, protocol supplementary concept word, rare disease supplementary concept word, unique identifier, synonyms]
11. (hand adj3 sanit*).mp. [mp=title, abstract, original title, name of substance word, subject heading word, floating sub-heading word, keyword heading word, organism supplementary concept word, protocol supplementary concept word, rare disease supplementary concept word, unique identifier, synonyms]
12. (hand adj3 disinfect*).mp. [mp=title, abstract, original title, name of substance word, subject heading word, floating sub-heading word, keyword heading word, organism supplementary concept word, protocol supplementary concept word, rare disease supplementary concept word, unique identifier, synonyms]
13. hand-sanit*.mp. [mp=title, abstract, original title, name of substance word, subject heading word, floating sub-heading word, keyword heading word, organism supplementary concept word, protocol supplementary concept word, rare disease supplementary concept word, unique identifier, synonyms]
14. (hand adj3 clean*).mp. [mp=title, abstract, original title, name of substance word, subject heading word, floating sub-heading word, keyword heading word, organism supplementary concept word, protocol supplementary concept word, rare disease supplementary concept word, unique identifier, synonyms]
15. hand-wash*.mp. [mp=title, abstract, original title, name of substance word, subject heading word, floating sub-heading word, keyword heading word, organism supplementary concept word, protocol supplementary concept word, rare disease supplementary concept word, unique identifier, synonyms]
16. hand-hygiene.mp. [mp=title, abstract, original title, name of substance word, subject heading word, floating sub-heading word, keyword heading word, organism supplementary concept word, protocol supplementary concept word, rare disease supplementary concept word, unique identifier, synonyms]
17. 8 or 9 or 10 or 11 or 12 or 13 or 14 or 15 or 16
18. exp Masks/
19. (face adj3 cover).mp. [mp=title, abstract, original title, name of substance word, subject heading word, floating sub-heading word, keyword heading word, organism supplementary concept word, protocol supplementary concept word, rare disease supplementary concept word, unique identifier, synonyms]
20. mask*.mp. [mp=title, abstract, original title, name of substance word, subject heading word, floating sub-heading word, keyword heading word, organism supplementary concept word, protocol supplementary concept word, rare disease supplementary concept word, unique identifier, synonyms]
21. (wear adj3 mask).mp. [mp=title, abstract, original title, name of substance word, subject heading word, floating sub-heading word, keyword heading word, organism supplementary concept word, protocol supplementary concept word, rare disease supplementary concept word, unique identifier, synonyms]
22. (facial adj3 cover*).mp. [mp=title, abstract, original title, name of substance word, subject heading word, floating sub-heading word, keyword heading word, organism supplementary concept word, protocol supplementary concept word, rare disease supplementary concept word, unique identifier, synonyms]
23. (facial adj3 mask).mp. [mp=title, abstract, original title, name of substance word, subject heading word, floating sub-heading word, keyword heading word, organism supplementary concept word, protocol supplementary concept word, rare disease supplementary concept word, unique identifier, synonyms]
24. facial-cover*.mp. [mp=title, abstract, original title, name of substance word, subject heading word, floating sub-heading word, keyword heading word, organism supplementary concept word, protocol supplementary concept word, rare disease supplementary concept word, unique identifier, synonyms]
25. (wore adj3 mask).mp. [mp=title, abstract, original title, name of substance word, subject heading word, floating sub-heading word, keyword heading word, organism supplementary concept word, protocol supplementary concept word, rare disease supplementary concept word, unique identifier, synonyms]
26. 18 or 19 or 20 or 21 or 22 or 23 or 24 or 25
27. exp Physical Distancing/
28. (social* adj3 distanc*).mp. [mp=title, abstract, original title, name of substance word, subject heading word, floating sub-heading word, keyword heading word, organism supplementary concept word, protocol supplementary concept word, rare disease supplementary concept word, unique identifier, synonyms]
29. (physical* adj3 distanc*).mp. [mp=title, abstract, original title, name of substance word, subject heading word, floating sub-heading word, keyword heading word, organism supplementary concept word, protocol supplementary concept word, rare disease supplementary concept word, unique identifier, synonyms]
30. (safe adj3 distanc*).mp. [mp=title, abstract, original title, name of substance word, subject heading word, floating sub-heading word, keyword heading word, organism supplementary concept word, protocol supplementary concept word, rare disease supplementary concept word, unique identifier, synonyms]
31. 27 or 28 or 29 or 30
32. exp Video Recording/
33. CCTV.mp. [mp=title, abstract, original title, name of substance word, subject heading word, floating sub-heading word, keyword heading word, organism supplementary concept word, protocol supplementary concept word, rare disease supplementary concept word, unique identifier, synonyms]
34. (closed adj3 circuit adj3 television).mp. [mp=title, abstract, original title, name of substance word, subject heading word, floating sub-heading word, keyword heading word, organism supplementary concept word, protocol supplementary concept word, rare disease supplementary concept word, unique identifier, synonyms]
35. (surveillance adj3 camera).mp. [mp=title, abstract, original title, name of substance word, subject heading word, floating sub-heading word, keyword heading word, organism supplementary concept word, protocol supplementary concept word, rare disease supplementary concept word, unique identifier, synonyms]
36. 32 or 33 or 34 or 35
37. exp Observational Study/
38. observ*.mp. [mp=title, abstract, original title, name of substance word, subject heading word, floating sub-heading word, keyword heading word, organism supplementary concept word, protocol supplementary concept word, rare disease supplementary concept word, unique identifier, synonyms]
39. 37 or 38
40. 17 or 26 or 31
41. 7 and 40
42. 39 and 41
43. 7 and 36 and 40
44. 42 or 43

SEARCH STRATEDGY Aim 3

1. exp hand washing/
2. (hand adj3 wash*).mp. [mp=title, abstract, heading word, drug trade name, original title, device manufacturer, drug manufacturer, device trade name, keyword, floating subheading word, candidate term word]
3. (hand adj3 hygiene).mp. [mp=title, abstract, heading word, drug trade name, original title, device manufacturer, drug manufacturer, device trade name, keyword, floating subheading word, candidate term word]
4. (hand adj3 sanit*).mp. [mp=title, abstract, heading word, drug trade name, original title, device manufacturer, drug manufacturer, device trade name, keyword, floating subheading word, candidate term word]
5. (hand adj3 disinect*).mp. [mp=title, abstract, heading word, drug trade name, original title, device manufacturer, drug manufacturer, device trade name, keyword, floating subheading word, candidate term word]
6. hand-sanit*.mp. [mp=title, abstract, heading word, drug trade name, original title, device manufacturer, drug manufacturer, device trade name, keyword, floating subheading word, candidate term word]
7. (hand adj3 clean*).mp. [mp=title, abstract, heading word, drug trade name, original title, device manufacturer, drug manufacturer, device trade name, keyword, floating subheading word, candidate term word]
8. hand-wash*.mp. [mp=title, abstract, heading word, drug trade name, original title, device manufacturer, drug manufacturer, device trade name, keyword, floating subheading word, candidate term word]
9. hand-hygiene.mp. [mp=title, abstract, heading word, drug trade name, original title, device manufacturer, drug manufacturer, device trade name, keyword, floating subheading word, candidate term word]
10. 1 or 2 or 3 or 4 or 5 or 6 or 7 or 8 or 9
11. exp face mask ventilation/ or exp mask/ or exp cloth mask/ or exp face mask/ or exp pediatric face mask/ or exp surgical mask/
12. (face adj3 cover*).mp. [mp=title, abstract, heading word, drug trade name, original title, device manufacturer, drug manufacturer, device trade name, keyword, floating subheading word, candidate term word]
13. mask*.mp.
14. (wear adj3 mask).mp. [mp=title, abstract, heading word, drug trade name, original title, device manufacturer, drug manufacturer, device trade name, keyword, floating subheading word, candidate term word]
15. (facial adj3 cover*).mp. [mp=title, abstract, heading word, drug trade name, original title, device manufacturer, drug manufacturer, device trade name, keyword, floating subheading word, candidate term word]
16. (facial adj3 mask).mp. [mp=title, abstract, heading word, drug trade name, original title, device manufacturer, drug manufacturer, device trade name, keyword, floating subheading word, candidate term word]
17. facial-cover*.mp. [mp=title, abstract, heading word, drug trade name, original title, device manufacturer, drug manufacturer, device trade name, keyword, floating subheading word, candidate term word]
18. (wore adj3 mask).mp. [mp=title, abstract, heading word, drug trade name, original title, device manufacturer, drug manufacturer, device trade name, keyword, floating subheading word, candidate term word]
19. 11 or 12 or 13 or 14 or 15 or 16 or 17 or 18
20. exp social distancing/
21. (social* adj3 distanc*).mp. [mp=title, abstract, heading word, drug trade name, original title, device manufacturer, drug manufacturer, device trade name, keyword, floating subheading word, candidate term word]
22. (physical* adj3 distanc*).mp. [mp=title, abstract, heading word, drug trade name, original title, device manufacturer, drug manufacturer, device trade name, keyword, floating subheading word, candidate term word]
23. (safe adj3 distanc*).mp. [mp=title, abstract, heading word, drug trade name, original title, device manufacturer, drug manufacturer, device trade name, keyword, floating subheading word, candidate term word]
24. 20 or 21 or 22 or 23
25. CCTV.mp.
26. (closed adj3 circuit adj3 television).mp. [mp=title, abstract, heading word, drug trade name, original title, device manufacturer, drug manufacturer, device trade name, keyword, floating subheading word, candidate term word]
27. exp videorecording/
28. (surveillance adj3 camera).mp. [mp=title, abstract, heading word, drug trade name, original title, device manufacturer, drug manufacturer, device trade name, keyword, floating subheading word, candidate term word]
29. 25 or 26 or 27 or 28
30. exp observational study/ or exp observational method/
31. observ*.mp. [mp=title, abstract, heading word, drug trade name, original title, device manufacturer, drug manufacturer, device trade name, keyword, floating subheading word, candidate term word]
32. exp infection/
33. 10 or 19 or 24
34. 32 and 33
35. 30 or 31
36. 29 or 35
37. 34 and 36

**Table 4:**
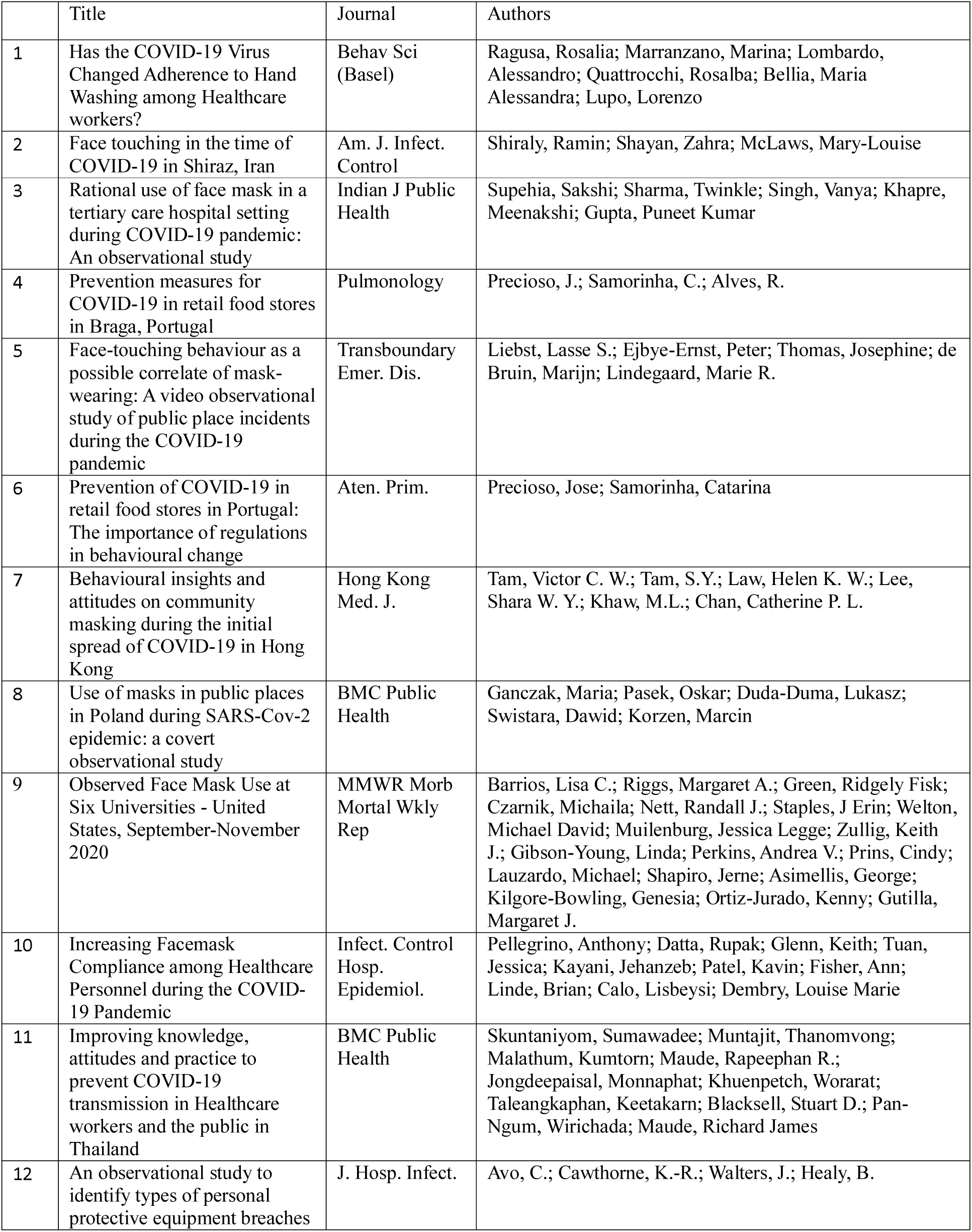
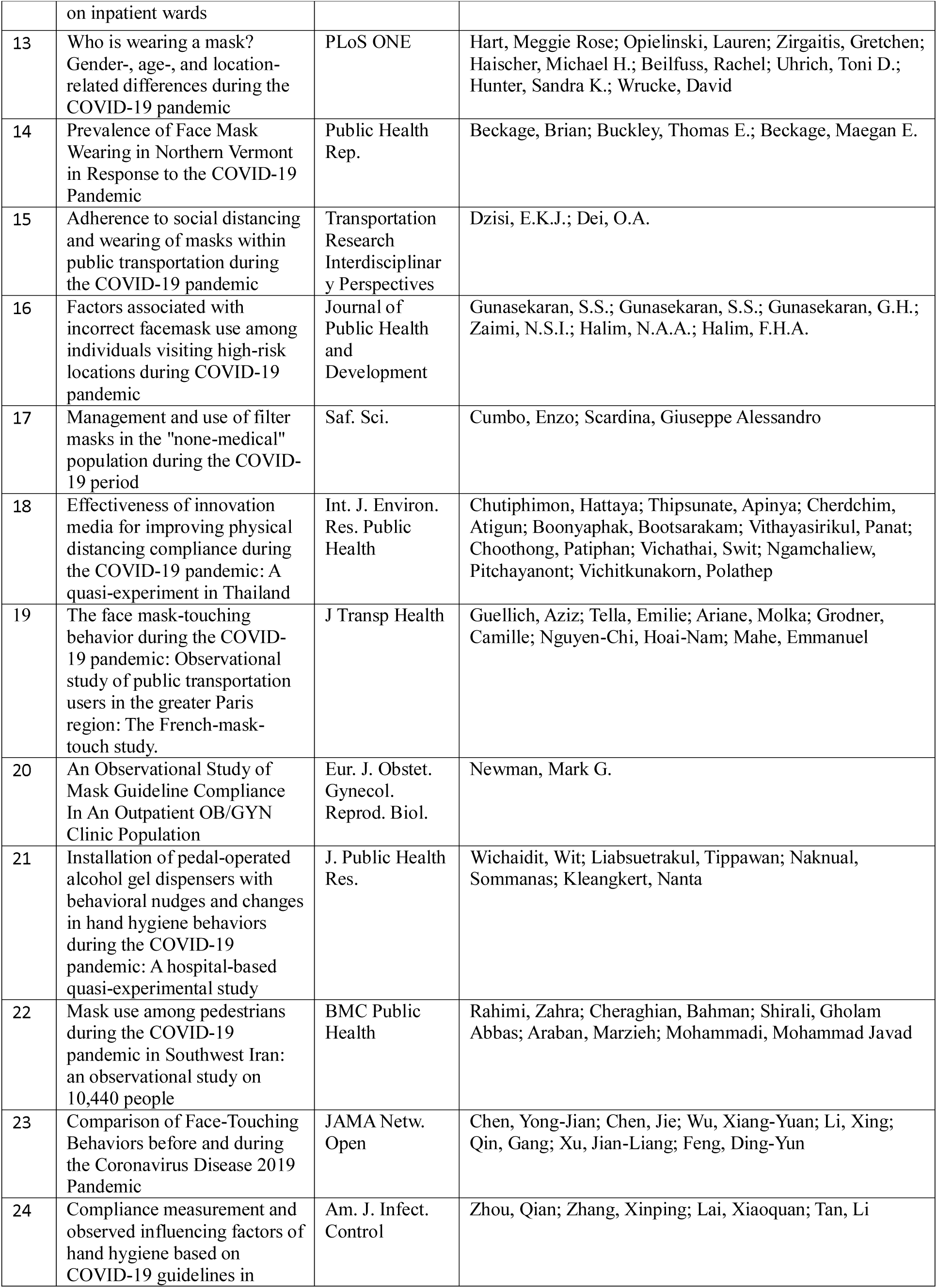
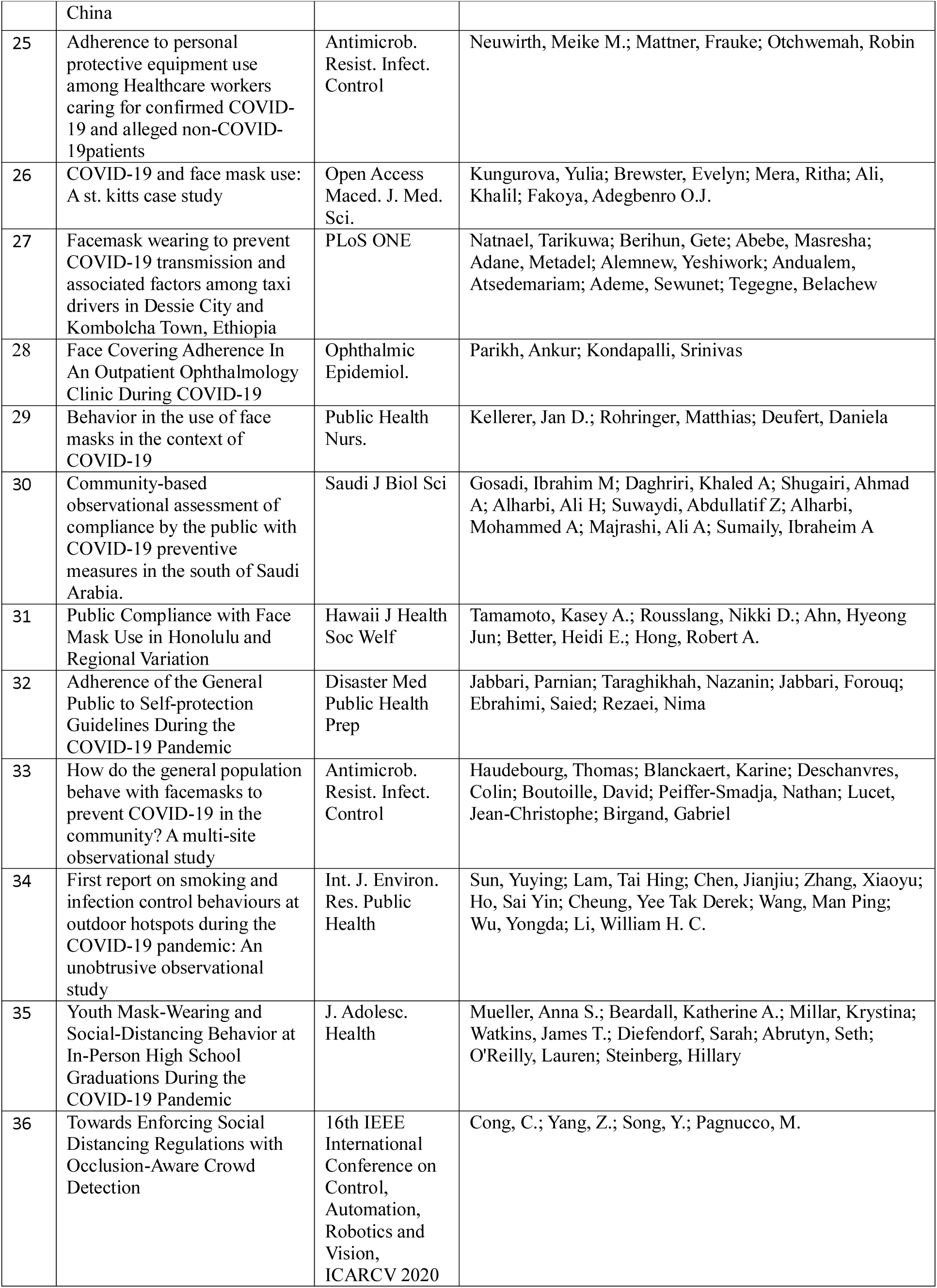
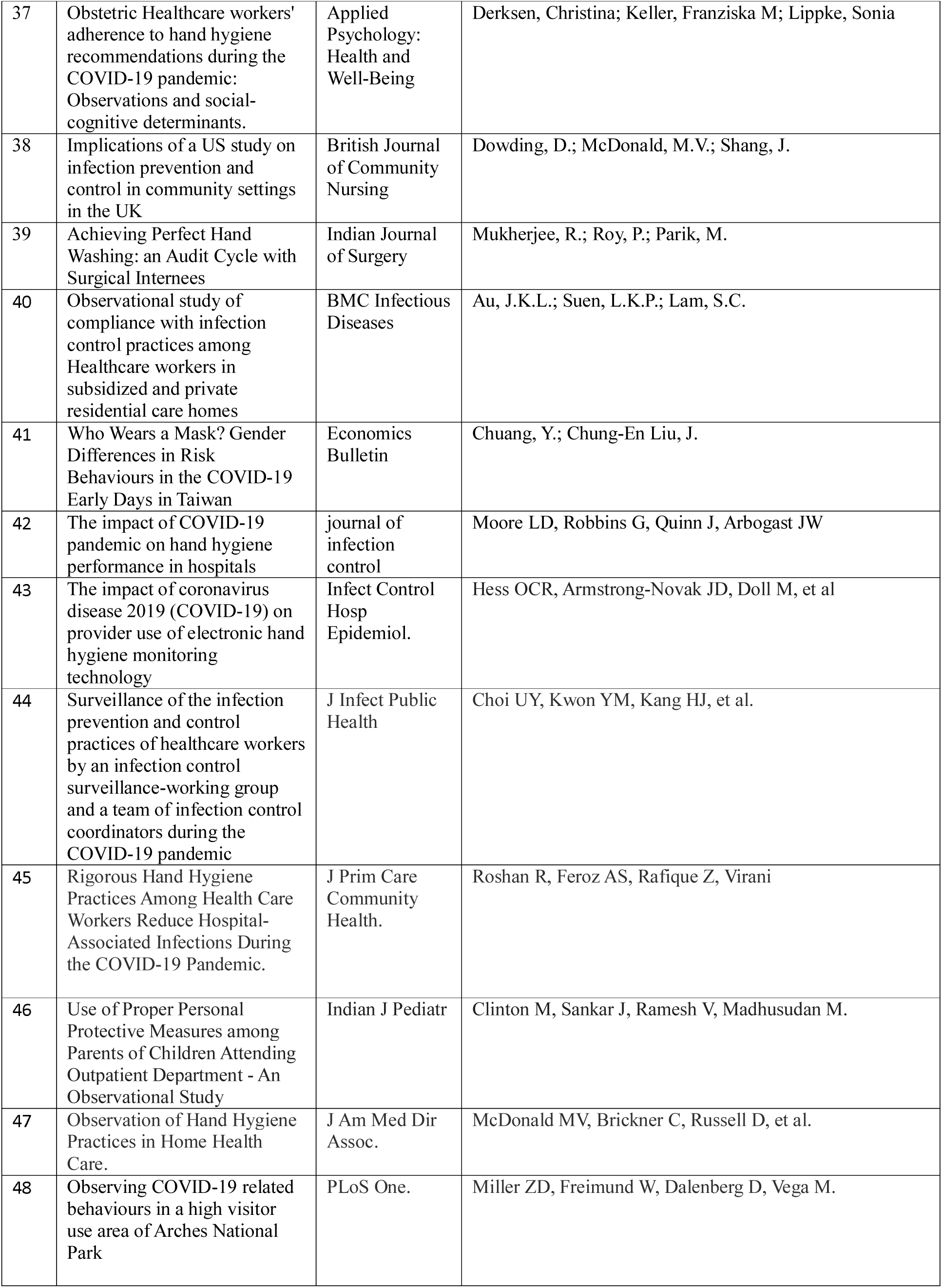
COVID-19 Included papers in data synthesis

**Table 5:**
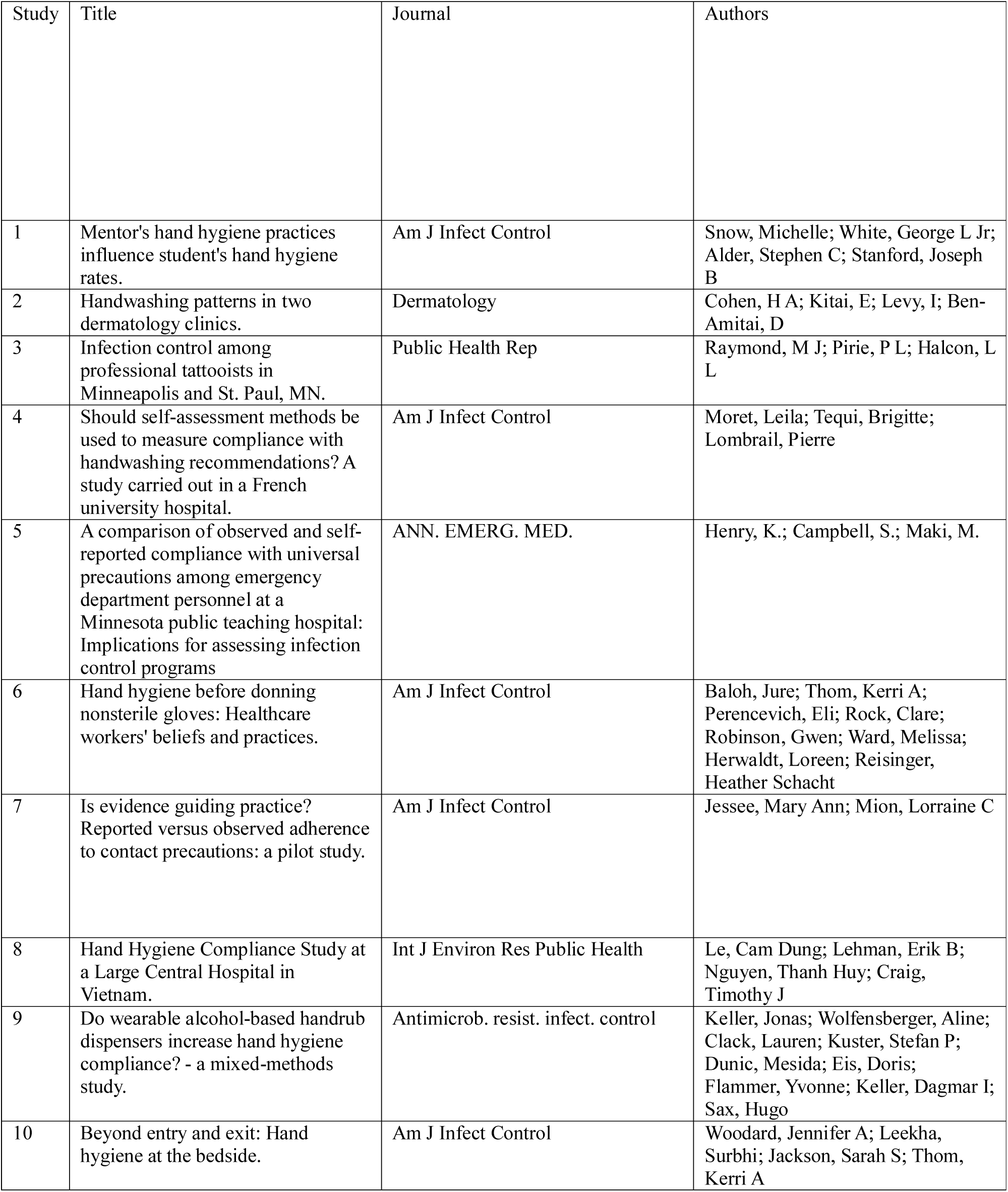
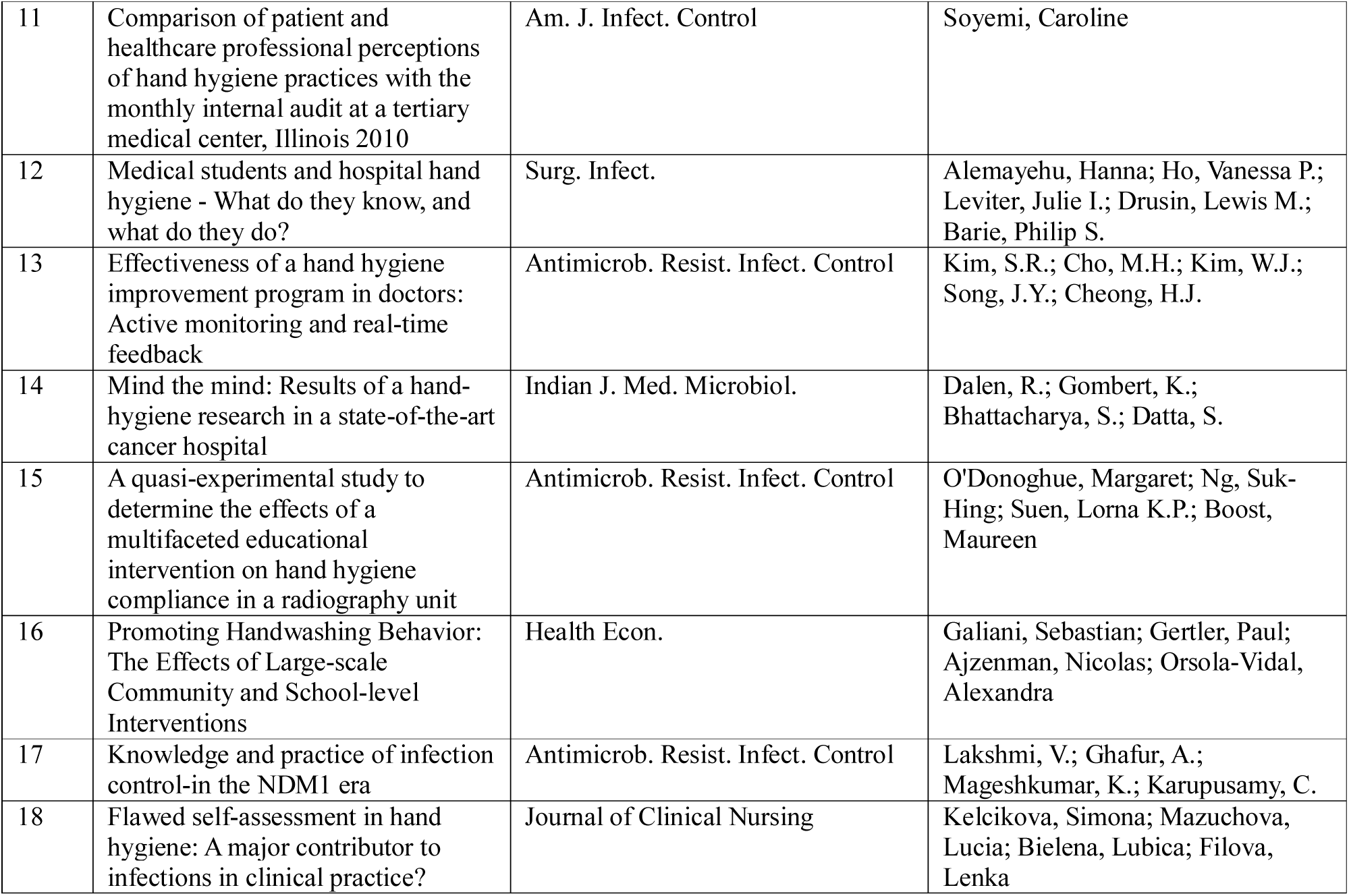
NON-COVID-19 papers included in synthesis

**Table 7:** NIH quality assessment checklist

| Criteria | Yes | No | Other<br>(CD,<br>NR,<br>NA)* |
| --- | --- | --- | --- |
| 1. Was the research question or objective in this paper clearly stated? |  |  |  |
| 2. Was the study population clearly specified and defined? |  |  |  |
| 3. Was the participation rate of eligible persons at least 50%? |  |  |  |
| 4. Were all the subjects selected or recruited from the same or similar populations (including the same time period)? Were inclusion and exclusion criteria for being in the study prespecified and applied uniformly to all participants? |  |  |  |
| 5. Was a sample size justification, power description, or variance and effect estimates provided? |  |  |  |
| 6. For the analyses in this paper, were the exposure(s) of interest measured prior to the outcome(s) being measured? |  |  |  |
| 7. Was the timeframe sufficient so that one could reasonably expect to see an association between exposure and outcome if it existed? |  |  |  |
| 8. For exposures that can vary in amount or level, did the study examine different levels of the exposure as related to the outcome (e.g., categories of exposure, or exposure measured as continuous variable)? |  |  |  |
| 9. Were the exposure measures (independent variables) clearly defined, valid, reliable, and implemented consistently across all study participants? |  |  |  |
| 10. Was the exposure(s) assessed more than once over time? |  |  |  |
| 11. Were the outcome measures (dependent variables) clearly defined, valid, reliable, and implemented consistently across all study participants? |  |  |  |
| 12. Were the outcome assessors blinded to the exposure status of participants? |  |  |  |
| 13. Was loss to follow-up after baseline 20% or less? |  |  |  |
| 14. Were key potential confounding variables measured and adjusted statistically for their impact on the relationship between exposure(s) and outcome(s)? |  |  |  |

## References

1. Scholz U, Freund AM. Determinants of protective behaviours during a nationwide lockdown in the wake of the COVID-19pandemic. Br J Health Psychol. 2021;26(3):935–957. doi:10.1111/bjhp.12513

2. Center for Disease Control. Social Distancing. Available at https://www.cdc.gov/coronavirus/2019-ncov/prevent-getting-sick/social-distancing.html (Accessed 09 June 2021)

3. https://www.bmj.com/content/375/bmj-2021-068302

4. Williams SN, Armitage CJ, Tampe T, Dienes KD. Public perceptions of non-adherence to COVID-19measures by self and others in the United Kingdom. MedRxiv [Preprint]. 2020;10.1101/2020.11.17.20233486

5. Tesfaye W, Peterson G. Self-reported medication adherence measurement tools: Some options to avoid a legal minefield [published online ahead of print, 2021 Aug 25]. J Clin Pharm Ther. 2021;10.1111/jcpt.13515. doi:10.1111/jcpt.13515

6. Mangtani P, Shah A, Roberts JA. Validation of influenza and pneumococcal vaccine status in adults based on self-report. Epidemiology and Infection. 2007;135(1):139–143. doi:10.1017/S0950268806006479

7. Kormos, C. & Gifford, R. The validity of self-report measures of proenvironmental behavior: A meta-analytic review. J. Environ. Psychol.2014. 40, 359–371

8. Dobbinson SJ, Jamsen K, Dixon HG, et al. Assessing population-wide behaviour change: concordance of 10-year trends in self-reported and observed sun protection. Int J Public Health. 2014;59(1):157–166. doi:10.1007/s00038-013-0454-5

9. Prince SA, Adamo KB, Hamel ME, Hardt J, Connor Gorber S, Tremblay M. A comparison of direct versus self-report measures for assessing physical activity in adults: a systematic review. Int J Behav Nutr Phys Act. 2008;5:56. Published 2008 Nov 6. doi:10.1186/1479-5868-5-56

10. Smith LE, Mottershaw AL, Egan M, Waller J, Marteau TM, Rubin GJ. Correction: The impact of believing you have had COVID-19on self-reported behaviour: Cross-sectional survey. PLoS One. 2021;16(2):e0248076. Published 2021 Feb 25. doi:10.1371/journal.pone.0248076

11. Jeffrey B, Walters CE, Ainslie KEC, et al. Anonymised and aggregated crowd level mobility data from mobile phones suggests that initial compliance with COVID-19social distancing interventions was high and geographically consistent across the UK. Wellcome Open Res. 2020;5:170. Published 2020 Jul 17. doi:10.12688/wellcomeopenres.15997.1

12. Chen L, Grimstead I, Bell D, et al. Estimating Vehicle and Pedestrian Activity from Town and City Traffic Cameras. Sensors (Basel). 2021;21(13):4564. Published 2021 Jul 3. doi:10.3390/s21134564

13. Glampson B, Brittain J, Kaura A, Mulla A, Mercuri L, Brett SJ, Aylin P, Sandall T, Goodman I, Redhead J, Saravanakumar K, Mayer EK. North West London COVID-19Vaccination Programme: Real-world evidence for Vaccine uptake and effectiveness: Retrospective Cohort Study. JMIR Public Health Surveill. 2021 Jul 8. doi: 10.2196/30010. Epub ahead of print. PMID: 34265740.

14. NIH. 2021. Study Quality Assessment Tool. Available at:https://www.nhlbi.nih.gov/health-topics/study-quality-assessment-tools (Accessed 1 October 2021).

15. Ragusa R, Marranzano M, Lombardo A, Quattrocchi R, Bellia MA, Lupo L. Has the COVID-19 19 Virus Changed Adherence to Hand Washing among Healthcare Workers?. Behav Sci (Basel). 2021;11(4):53. Published 2021 Apr 15. doi:10.3390/bs11040053

16. Shiraly R, Shayan Z, McLaws ML. Face touching in the time of COVID-19in Shiraz, Iran. Am J Infect Control. 2020;48(12):1559–1561. doi:10.1016/j.ajic.2020.08.009

17. Supehia S, Singh V, Sharma T, Khapre M, Gupta PK. Rational use of face mask in a tertiary care hospital setting during COVID-19pandemic: An observational study. Indian J Public Health. 2020;64(Supplement):S225–S227. doi:10.4103/ijph.IJPH_493_20

18. Precioso J, Samorinha C, Alves R. Prevention measures for COVID-19in retail food stores in Braga, Portugal. Pulmonology. 2021;27(3):260–261. doi:10.1016/j.pulmoe.2020.06.009

19. Liebst LS, Ejbye-Ernst P, de Bruin M, Thomas J, Lindegaard MR. Face-touching behaviour as a possible correlate of mask-wearing: A video observational study of public place incidents during the COVID-19pandemic [published online ahead of print, 2021 Apr 5]. Transbound Emerg Dis. 2021;10.1111/tbed.14094. doi:10.1111/tbed.14094

20. Precioso J, Samorinha C. Prevention of COVID-19in retail food stores in Portugal: The importance of regulations in behavioural change. Aten Primaria. 2021;53(2):101953. doi:10.1016/j.aprim.2020.07.011

21. Tam VCW, Tam SY, Khaw ML, Law HKW, Chan CPL, Lee SWY. Behavioural insights and attitudes on community masking during the initial spread of COVID-19in Hong Kong. Hong Kong Med J. 2021;27(2):106–112. doi:10.12809/hkmj209015

22. Ganczak M, Pasek O, Duda-Duma Ł, Świstara D, Korzeń M. Use of masks in public places in Poland during SARS-Cov-2 epidemic: a covert observational study. BMC Public Health. 2021;21(1):393. Published 2021 Feb 23. doi:10.1186/s12889-021-10418-3

23. Barrios LC, Riggs MA, Green RF, et al. Observed Face Mask Use at Six Universities - United States, September-November 2020. MMWR Morb Mortal Wkly Rep. 2021;70(6):208–211. Published 2021 Feb 12. doi:10.15585/mmwr.mm7006e1

24. Datta R, Glenn K, Pellegrino A, et al. Increasing face-mask compliance among healthcare personnel during the coronavirus disease 2019 (COVID-19-19) pandemic [published online ahead of print, 2021 May 3]. Infect Control Hosp Epidemiol. 2021;1–7. doi:10.1017/ice.2021.205

25. Maude RR, Jongdeepaisal M, Skuntaniyom S, et al. Improving knowledge, attitudes and practice to prevent COVID-19transmission in healthcare workers and the public in Thailand. BMC Public Health. 2021;21(1):749. Published 2021 Apr 18. doi:10.1186/s12889-021-10768-y

26. Avo C, Cawthorne KR, Walters J, Healy B. An observational study to identify types of personal protective equipment breaches on inpatient wards. J Hosp Infect. 2020;106(1):208–210. doi:10.1016/j.jhin.2020.06.024

27. Haischer MH, Beilfuss R, Hart MR, et al. Who is wearing a mask? Gender-, age-, and location-related differences during the COVID-19pandemic. PLoS One. 2020;15(10):e0240785. Published 2020 Oct 15. doi:10.1371/journal.pone.0240785

28. Beckage B, Buckley TE, Beckage ME. Prevalence of Face Mask Wearing in Northern Vermont in Response to the COVID-19Pandemic. Public Health Rep. 2021;136(4):451–456. doi:10.1177/00333549211009496

29. Dzisi EKJ, Dei OA. Adherence to social distancing and wearing of masks within public transportation during the COVID-19 19 pandemic. Transp Res Interdiscip Perspect. 2020;7:100191. doi:10.1016/j.trip.2020.100191

30. Gunasekaran, S.S.; Gunasekaran, S.S.; Gunasekaran, G.H.; Zaimi, N.S.I.; Halim, N.A.A.; Halim, F.H.A. Factors associated with incorrect facemask use among individuals visiting high-risk locations during COVID-19pandemic. Int. J. Public Health 2020, 18, 38–48.

31. Cumbo E, Scardina GA. Management and use of filter masks in the “none-medical” population during the COVID-19period. Saf Sci. 2021;133:104997. doi:10.1016/j.ssci.2020.104997

32. Chutiphimon H, Thipsunate A, Cherdchim A, et al. Effectiveness of Innovation Media for Improving Physical Distancing Compliance during the COVID-19Pandemic: A Quasi-Experiment in Thailand. Int J Environ Res Public Health. 2020;17(22):8535. Published 2020 Nov 17. doi:10.3390/ijerph17228535

33. Guellich A, Tella E, Ariane M, Grodner C, Nguyen-Chi HN, Mahé E. The face mask-touching behavior during the COVID-19pandemic: Observational study of public transportation users in the greater Paris region: The French-mask-touch study. J Transp Health. 2021;21:101078. doi:10.1016/j.jth.2021.101078

34. Newman MG. An Observational Study of Mask Guideline Compliance In An Outpatient OB/GYN Clinic Population. Eur J Obstet Gynecol Reprod Biol. 2020;255:268–269. doi:10.1016/j.ejogrb.2020.10.048

35. Wichaidit W, Naknual S, Kleangkert N, Liabsuetrakul T. Installation of pedal-operated alcohol gel dispensers with behavioral nudges and changes in hand hygiene behaviors during the COVID-19pandemic: A hospital-based quasi-experimental study. J Public Health Res. 2020;9(4):1863. Published 2020 Oct 26. doi:10.4081/jphr.2020.1863

36. Rahimi Z, Shirali GA, Araban M, Mohammadi MJ, Cheraghian B. Mask use among pedestrians during the COVID-19pandemic in Southwest Iran: an observational study on 10,440 people. BMC Public Health. 2021;21(1):133. Published 2021 Jan 14. doi:10.1186/s12889-020-10152-2

37. Chen YJ, Qin G, Chen J, et al. Comparison of Face-Touching Behaviors Before and During the Coronavirus Disease 2019 Pandemic. JAMA Netw Open. 2020;3(7):e2016924. Published 2020 Jul 1. doi:10.1001/jamanetworkopen.2020.16924

38. Zhou Q, Lai X, Zhang X, Tan L. Compliance measurement and observed influencing factors of hand hygiene based on COVID-19guidelines in China. Am J Infect Control. 2020;48(9):1074–1079. doi:10.1016/j.ajic.2020.05.043

39. Neuwirth MM, Mattner F, Otchwemah R. Adherence to personal protective equipment use among healthcare workers caring for confirmed COVID-19and alleged non-COVID-19patients. Antimicrob Resist Infect Control. 2020;9(1):199. Published 2020 Dec 10. doi:10.1186/s13756-020-00864-w

40. Kungurova, Y., Mera, R., Brewster, E., Ali, K., & Fakoya, A. O. (2020). COVID-19and face mask use: A St. kitts case study. Open access maced. Journal of Medical Sciences., 8, 346– 352. https://doi.org/10.17533/udea.iee.v38n2e13

41. Natnael T, Alemnew Y, Berihun G, et al. Facemask wearing to prevent COVID-19transmission and associated factors among taxi drivers in Dessie City and Kombolcha Town, Ethiopia. PLoS One. 2021;16(3):e0247954. Published 2021 Mar 12. doi:10.1371/journal.pone.0247954

42. Parikh A, Kondapalli S. FACE COVERING ADHERENCE IN AN OUTPATIENT OPHTHALMOLOGY CLINIC DURING COVID-19-19. Ophthalmic Epidemiol. 2021;28(4):365–368. doi:10.1080/09286586.2020.1866022

43. Kellerer JD, Rohringer M, Deufert D. Behavior in the use of face masks in the context of COVID-19-19. Public Health Nurs. 2021;38(5):862–868. doi:10.1111/phn.12918

44. Gosadi IM, Daghriri KA, Shugairi AA, et al. Community-based observational assessment of compliance by the public with COVID-1919 preventive measures in the south of Saudi Arabia. Saudi J Biol Sci. 2021;28(3):1938–1943. doi:10.1016/j.sjbs.2020.12.045

45. Tamamoto KA, Rousslang ND, Ahn HJ, Better HE, Hong RA. Public Compliance with Face Mask Use in Honolulu and Regional Variation. Hawaii J Health Soc Welf. 2020;79(9):268–271.

46. Jabbari P, Taraghikhah N, Jabbari F, Ebrahimi S, Rezaei N. Adherence of the General Public to Self-Protection Guidelines During the COVID-19Pandemic [published online ahead of print, 2020 Nov 18]. Disaster Med Public Health Prep. 2020;1–4. doi:10.1017/dmp.2020.445

47. Deschanvres C, Haudebourg T, Peiffer-Smadja N, et al. How do the general population behave with facemasks to prevent COVID-19in the community? A multi-site observational study. Antimicrob Resist Infect Control. 2021;10(1):61. Published 2021 Mar 29. doi:10.1186/s13756-021-00927-6

48. Sun Y, Lam TH, Cheung YTD, et al. First Report on Smoking and Infection Control Behaviours at Outdoor Hotspots during the COVID-19Pandemic: An Unobtrusive Observational Study. Int J Environ Res Public Health. 2021;18(3):1031. Published 2021 Jan 25. doi:10.3390/ijerph18031031

49. Mueller AS, Diefendorf S, Abrutyn S, et al. Youth Mask-Wearing and Social-Distancing Behavior at In-Person High School Graduations During the COVID-19Pandemic. J Adolesc Health. 2021;68(3):464–471. doi:10.1016/j.jadohealth.2020.12.123

50. C. Cong, Z. Yang, Y. Song and M. Pagnucco, “Towards Enforcing Social Distancing Regulations with Occlusion-Aware Crowd Detection,” 2020 16th International Conference on Control, Automation, Robotics and Vision (ICARCV), 2020, pp. 297–302, doi: 10.1109/ICARCV50220.2020.9305507.

51. Derksen C, Keller FM, Lippke S. Obstetric Healthcare Workers’ Adherence to Hand Hygiene Recommendations during the COVID-19Pandemic: Observations and Social-Cognitive Determinants. Appl Psychol Health Well Being. 2020;12(4):1286–1305. doi:10.1111/aphw.12240

52. Dowding D, McDonald MV, Shang J. Implications of a US study on infection prevention and control in community settings in the UK. Br J Community Nurs. 2020;25(12):578–583. doi:10.12968/bjcn.2020.25.12.578

53. Mukherjee R, Roy P, Parik M. Achieving Perfect Hand Washing: an Audit Cycle with Surgical Internees [published online ahead of print, 2020 Oct 6]. Indian J Surg. 2020;1–7. doi:10.1007/s12262-020-02619-8

54. Au JKL, Suen LKP, Lam SC. Observational study of compliance with infection control practices among healthcare workers in subsidized and private residential care homes. BMC Infect Dis. 2021;21(1):75. Published 2021 Jan 14. doi:10.1186/s12879-021-05767-8

55. Chuang, Y., and Liu, J. C. E. (2020). Who wears a mask? Gender differences in risk behaviors in the COVID-19early days in Taiwan. Econ. Bull. 40, 2619–2627. Available online at: https://ideas.repec.org/a/ebl/ecbull/eb-20-00882.html

56. Moore LD, Robbins G, Quinn J, Arbogast JW. The impact of COVID-19pandemic on hand hygiene performance in hospitals. Am J Infect Control. 2021;49(1):30–33. doi:10.1016/j.ajic.2020.08.021

57. Hess OCR, Armstrong-Novak JD, Doll M, et al. The impact of coronavirus disease 2019 (COVID-19-19) on provider use of electronic hand hygiene monitoring technology. Infect Control Hosp Epidemiol. 2021;42(8):1007–1009. doi:10.1017/ice.2020.1336

58. Choi UY, Kwon YM, Kang HJ, et al. Surveillance of the infection prevention and control practices of healthcare workers by an infection control surveillance-working group and a team of infection control coordinators during the COVID-19pandemic. J Infect Public Health. 2021;14(4):454–460. doi:10.1016/j.jiph.2021.01.012

59. Roshan R, Feroz AS, Rafique Z, Virani N. Rigorous Hand Hygiene Practices Among Health Care Workers Reduce Hospital-Associated Infections During the COVID-19Pandemic. J Prim Care Community Health. 2020;11:2150132720943331. doi:10.1177/2150132720943331

60. Clinton M, Sankar J, Ramesh V, Madhusudan M. Use of Proper Personal Protective Measures among Parents of Children Attending Outpatient Department - An Observational Study. Indian J Pediatr. 2021;88(5):480. doi:10.1007/s12098-020-03624-1

61. McDonald MV, Brickner C, Russell D, et al. Observation of Hand Hygiene Practices in Home Health Care. J Am Med Dir Assoc. 2021;22(5):1029–1034. doi:10.1016/j.jamda.2020.07.031

62. Miller ZD, Freimund W, Dalenberg D, Vega M. Observing COVID-19related behaviors in a high visitor use area of Arches National Park. PLoS One. 2021;16(2):e0247315. Published 2021 Feb 22. doi:10.1371/journal.pone.0247315

63. Snow M, White GL Jr, Alder SC, Stanford JB. Mentor’s hand hygiene practices influence student’s hand hygiene rates. Am J Infect Control. 2006;34(1):18–24. doi:10.1016/j.ajic.2005.05.009

64. Cohen HA, Kitai E, Levy I, Ben-Amitai D. Handwashing patterns in two dermatology clinics. Dermatology. 2002;205(4):358–361. doi:10.1159/000066421

65. Raymond MJ, Pirie PL, Halcón LL. Infection control among professional tattooists in Minneapolis and St. Paul, MN. Public Health Rep. 2001;116(3):249–256. doi:10.1093/phr/116.3.249

66. Moret L, Tequi B, Lombrail P. Should self-assessment methods be used to measure compliance with handwashing recommendations? A study carried out in a French university hospital. Am J Infect Control. 2004;32(7):384–390. doi:10.1016/j.ajic.2004.02.004

67. Henry K, Campbell S, Maki M. A comparison of observed and self-reported compliance with universal precautions among emergency department personnel at a Minnesota public teaching hospital: implications for assessing infection control programs. Ann Emerg Med. 1992;21(8):940–946. doi:10.1016/s0196-0644(05)82932-4

68. Baloh J, Thom KA, Perencevich E, et al. Hand hygiene before donning nonsterile gloves: Healthcareworkers’ beliefs and practices. Am J Infect Control. 2019;47(5):492–497. doi:10.1016/j.ajic.2018.11.015

69. Jessee MA, Mion LC. Is evidence guiding practice? Reported versus observed adherence to contact precautions: a pilot study. Am J Infect Control. 2013;41(11):965–970. doi:10.1016/j.ajic.2013.05.005

70. Le CD, Lehman EB, Nguyen TH, Craig TJ. Hand Hygiene Compliance Study at a Large Central Hospital in Vietnam. Int J Environ Res Public Health. 2019;16(4):607. Published 2019 Feb 19. doi:10.3390/ijerph16040607

71. Keller J, Wolfensberger A, Clack L, et al. Do wearable alcohol-based handrub dispensers increase hand hygiene compliance? - a mixed-methods study. Antimicrob Resist Infect Control. 2018;7:143. Published 2018 Nov 23. doi:10.1186/s13756-018-0439-5

72. Woodard JA, Leekha S, Jackson SS, Thom KA. Beyond entry and exit: Hand hygiene at the bedside. Am J Infect Control. 2019;47(5):487–491. doi:10.1016/j.ajic.2018.10.026

73. C Soyemi et.al.Comparison of Patient and Healthcare Professional Perceptions of Hand Hygiene Practices with the Monthly Internal Audit at a Tertiary Medical Center. American Journal of Infection Control.2011;39(5)

74. Alemayehu H, Ho V et al. Medical students and hospital hand hygiene - What do they know, and what do they do? P. Surg. Infect. 2011, 12: S1

75. Kim, S., Cho, M., Kim, W. et al. P137: Effectiveness of a hand hygiene improvement program in doctors: active monitoring and real-time feedback. Antimicrob Resist Infect Control 2, P137 (2013). https://doi.org/10.1186/2047-2994-2-S1-P137

76. van Dalen R, Gombert K, Bhattacharya S, Datta SS. Mind the mind: results of a hand-hygiene research in a state-of-the-art cancer hospital. Indian J Med Microbiol. 2013;31(3):280–282. doi:10.4103/0255-0857.115639

77. O’Donoghue M, Ng SH, Suen LK, Boost M. A quasi-experimental study to determine the effects of a multifaceted educational intervention on hand hygiene compliance in a radiography unit. Antimicrob Resist Infect Control. 2016;5:36. Published 2016 Oct 19. doi:10.1186/s13756-016-0133-4

78. Galiani S, Gertler P, Ajzenman N, Orsola-Vidal A. Promoting Handwashing Behavior: The Effects of Large-scale Community and School-level Interventions. Health Econ. 2016;25(12):1545–1559. doi:10.1002/hec.3273

79. Lakshmi, V., Ghafur, A., Mageshkumar, K. et al. Knowledge and practice of infection control – in the NDM1 era. Antimicrob Resist Infect Control 4, P118 (2015). https://doi.org/10.1186/2047-2994-4-S1-P118

80. Kelcikova S, Mazuchova L, Bielena L, Filova L. Flawed self-assessment in hand hygiene: A major contributor to infections in clinical practice?. J Clin Nurs. 2019;28(11-12):2265–2275. doi:10.1111/jocn.14823

81. Davies R et.al 1The impact of “freedom day”on COVID-19 health protective behaviour in England: An observational study of hand hygiene, face covering use and physical distancing in public spaces pre and post the relaxing of restrictions. OSF. Available from: OSF | Manuscript observational study - preprint 19-10-2021.pdf

82. Davies R, Weinman J, Rubin GJ. Observed and self-reported COVID-19 health protection behaviours on a university campus and the impact of a single simple intervention. MedRXiv. Available from: https://www.medrxiv.org/content/10.1101/2021.06.15.21258920v1

83. (Kormos, C. & Gifford, R. The validity of self - report measures of proenvironmental behavior: A meta - analytic review. J. Environ. Psychol. 2014. 40, 359 – 371)

84. Smith, L. E., Potts, H., Amlôt, R., Fear, N. T., Michie, S., & Rubin, G. J. (2021). Adherence to the test, trace, and isolate system in the UK: results from 37 nationally representative surveys. BMJ (Clinical research ed.), 372, n608. https://doi.org/10.1136/bmj.n608

85. www.ons.gov.uk. (2021). Coronavirus and self-isolation after testing positive in England - Office for National Statistics. [online] Available at: https://www.ons.gov.uk/peoplepopulationandcommunity/healthandsocialcare/healthandwellbeing/bulletins/coronavirusandselfisolationaftertestingpositiveinengland/27septemberto2october2021 [Accessed 19 Nov. 2021].

86. Mangtani, P., Shah, A., & Roberts, J. A. (2007). Validation of influenza and pneumococcal vaccine status in adults based on self-report. Epidemiology and infection, 135(1), 139–143. https://doi.org/10.1017/S0950268806006479

87. Rolnick, S. J., Parker, E. D., Nordin, J. D., Hedblom, B. D., Wei, F., Kerby, T., Jackson, J. M., Crain, A. L., & Euler, G. (2013). Self-report compared to electronic medical record across eight adult vaccines: do results vary by demographic factors?. Vaccine, 31(37), 3928–3935. https://doi.org/10.1016/j.vaccine.2013.06.041

88. Gupta, C et.al.Coronamask: A Face Mask Detector for Real-Time Data. International Journal of Advanced Trends in Computer Science and Engineering.2020;9. 5624–5630. 10.30534/ijatcse/2020/212942020.

89. Pouw CAS, Toschi F, van Schadewijk F, Corbetta A. Monitoring physical distancing for crowd management: Real-time trajectory and group analysis. PLoS One. 2020;29;15(10):e0240963. doi: 10.1371/journal.pone.0240963.

90. Cawthorne KR, Oliver C, Cooke RPD. A user’s view of commercial mobile applications designed to measure hand hygiene compliance by direct observation [published online ahead of print, 2021 Aug 14]. J Hosp Infect. 2021;117:4–8. doi:10.1016/j.jhin.2021.08.008

